# Real-World Twelve-Month Weight Loss Outcomes in a Comprehensive Obesity Care Program Utilizing Individualized Nutrition Therapy and Incretin-Based Obesity Medications

**DOI:** 10.64898/2026.10.09.26365158

**Authors:** Shaminie J Athinarayanan, Alison R. Zoller, Kristin E. Bergethon, Amy M Goss, Barbara A. Gower

## Abstract

**Background/Objectives:** Incretin-based obesity medications (IOMs) produce substantial weight loss, but the added benefit of structured nutritional therapy remains unclear. We compared 12-month weight loss between individualized carbohydrate-reduced nutritional therapy plus IOMs (VINT+IOM) and IOM-based care without structured nutritional therapy (IOM only).

**Methods:** In this retrospective cohort study of 4,269 adults in a telemedicine obesity care program (VINT+IOM, n = 1,658; IOM only, n = 2,611), both pathways included clinician-guided medication management and behavioral coaching. The primary analysis included GLP-1–naïve participants on a single ingredient (semaglutide or tirzepa ide). Adjusted weight change was estimated using longitudinal linear mixed-effects models.

**Results:** Retention through 12 months was 77% (VINT+IOM) vs. 72% (IOM only). In GLP-1–naïve semaglutide users, adjusted 12-month weight loss was greater with VINT+IOM than IOM only (13.9 vs. 10.9 kg; difference 3.0 kg, 95% CI 0.6–5.4; P = 0.012); observed loss was 13.3% vs. 11.1%. In tirzepatide users, adjusted loss was 20.8 vs. 15.6 kg (difference 5.2 kg, 95% CI 1.8–8.7; P = 0.003); observed loss was 19.7% vs. 15.6%. Responder rates favored VINT+IOM. In the overall cohort, adjusted loss was 13.3 vs. 9.8 kg (P < 0.001), and the advantage persisted among participants with prior GLP-1 exposure. Weight loss continued between 6 and 12 months, and approximately 80% of participants in both pathways had IOM proportion of days covered ≥80%. Self-reported side effects were similar (63.6% vs 64.6%).

**Conclusions:** Adding individualized carbohydrate-reduced nutritional therapy to IOM-based care was associated with greater 12-month weight loss than IOM-based care without structured nutritional therapy. These real-world findings support further evaluation of individualized nutrition as an adjunct to incretin-based pharmacotherapy.

## Introduction

Incretin-based obesity medications (IOMs), particularly glucagon-like peptide-1 receptor agonists (GLP-1 RAs) and dual glucose-dependent insulinotropic polypeptide (GIP)/GLP-1 RAs, have transformed obesity treatment by producing substantial weight loss and improvements in cardiometabolic health. However, achieving and sustaining clinically meaningful weight loss in routine clinical practice remains challenging, underscoring the need to identify approaches that enhance the effectiveness of IOM–based therapies. Real-world 12-month persistence and adherence to GLP-1–based therapies have historically been suboptimal, with earlier studies reporting persistence rates of 24.5%– 32.3%, mean proportion of days covered (PDC) of 51.0%–54.3%, and only 27.2%–29.6% of patients achieving PDC ≥80% [1,2]. However, more recent evidence from a retrospective claims-based cohort of commercially insured adults without diabetes who initiated weight loss–indicated GLP-1–based therapies in 2024 reported higher 12-month persistence rates of 58.6% for semaglutide and 64.8% for tirzepatide [3]. Nevertheless, medication switching, treatment interruptions, dose changes, and discontinuation are common and often unavoidable features of real-world obesity pharmacotherapy, underscoring the need for care models that can accommodate these dynamic treatment trajectories while providing continuity of support and individualized management [2,4]. The ability to navigate these treatment challenges is particularly important given the substantial and relatively rapid weight regain that can occur following medication discontinuation, estimated at approximately 0.5 kg per month overall and up to 0.8 kg per month following discontinuation of newer incretin therapies [5,6].

Digital health offers a scalable approach for integrating IOM pharmacotherapy with longitudinal clinical monitoring, medication management, behavioral support, and health coaching. Emerging evidence supports the integration of obesity pharmacotherapy with digitally delivered behavioral and lifestyle interventions, including app-based self-monitoring, remote clinical oversight, and individualized coaching [7]. Real-world studies of digitally delivered obesity care have demonstrated substantial weight loss among individuals receiving semaglutide or tirzepatide within these multimodal care models [7,8,9,10]. These findings derive largely from small observational cohorts and service evaluations without comparison groups, making it difficult to separate the contribution of digital support from that of pharmacotherapy itself, and, more importantly, to isolate the contribution of any specific component of care, such as nutrition.

Beyond digital delivery, lifestyle modification remains an important component of comprehensive obesity management alongside IOM pharmacotherapy, with potential contributions extending beyond weight loss alone. Nutritional intervention, physical activity, and behavioral support can address dietary quality, physical function, body composition, and behaviors relevant to long-term weight management. A recent systematic review and meta-analysis of 33 randomized trials involving 12,028 participants demonstrated that GLP-1 RAs delivered alongside lifestyle modification produced substantial improvements in body weight, waist circumference, glycemic measures, blood pressure, and lipid parameters [11]. Lifestyle intervention may also be particularly relevant to the quality of weight loss during IOM treatment, as substantial weight reduction can include loss of lean mass; a 2026 systematic review and meta-analysis found that incorporating resistance training and adequate protein intake may help preserve lean mass during weight-loss treatment [12]. Consistent with this evidence, contemporary clinical guidelines recommend that GLP-1–based pharmacotherapy be delivered alongside dietary, physical activity, and behavioral interventions [13,14]. However, this evidence has important limitations. Trials and meta-analyses of GLP-1 RAs with lifestyle modification generally evaluate broad, multicomponent lifestyle support rather than a defined nutritional strategy, and they typically compare medication plus lifestyle with medication alone or placebo, not one nutritional approach against another or against usual medication care within the same program [11]. Consequently, it remains unknown whether a structured, individualized nutritional therapy such as carbohydrate reduction adds additional cardiometabolic benefits beyond IOM treatment delivered with standard coaching and clinical support, particularly in real-world settings where dose changes, switching, interruptions, and prior GLP-1 exposure are common.

Virta’s telemedicine-based continuous remote care model has demonstrated sustained improvements in glycemic control, weight, and medication use among adults with type 2 diabetes [15,16] and has subsequently been extended to comprehensive obesity care integrating remote clinical oversight, health coaching, behavioral support, medication management, and individualized nutrition. Within this model, participants receiving IOMs may engage in either Virta Individualized Nutrition Therapy (VINT), which emphasizes personalized carbohydrate reduction with continuous monitoring, or IOM only intervention delivered within the same multidisciplinary care infrastructure. Although VINT has demonstrated effectiveness as a standalone intervention for overweight and obesity [17–19], to our knowledge, its incremental association with weight loss when combined with IOMs has not been evaluated. Because the VINT+IOM and IOM-only pathways share digital delivery, clinical oversight, coaching, and medication access while differing primarily in the intensity and individualization of nutritional therapy, they provide an uncommon opportunity to examine the added value of nutritional therapy under routine clinical conditions, with the care infrastructure held largely constant.

The primary objective was to compare 12-month weight loss between VINT+IOM and IOM-only among GLP-1–naïve participants receiving a single GLP-1–based active pharmaceutical ingredient (semaglutide or tirzepatide) during follow-up and remaining engaged in a single prescription pathway for at least 9 months. This analysis was intended to evaluate the additional weight loss associated with integrating carbohydrate-reduced nutritional therapy with GLP-1–based pharmacotherapy compared with pharmacotherapy alone. Ancillary analyses examined weight-loss outcomes in the overall prescription cohort and across additional medication exposure subgroups, including the broader GLP-1–naïve population, participants with prior GLP-1 exposure, and those who switched medication ingredients during follow-up. Among participants with prior GLP-1 exposure, cumulative weight loss from the pre-GLP-1 reference weight was also evaluated to contextualize additional reductions achieved during program participation. Longitudinal weight trajectories were examined to characterize the pattern of weight reduction over 12 months, including changes between 6 and 12 months. Secondary objectives included characterizing medication utilization patterns, dose distribution and escalation, medication switching, adherence, and reported side effects, as well as exploring associations between medication use patterns and weight-loss outcomes, and changes in HbA1c as an exploratory outcome.

This study advances existing knowledge in three ways. First, it compares individualized carbohydrate-reduced nutritional therapy plus IOM with IOM alone within a single care infrastructure, isolating nutrition support as the main difference between pathways, although residual confounding cannot be excluded. Second, it characterizes real-world medication patterns, including ingredient switching and prior GLP-1 exposure, and relates them to weight outcomes. These factors are typically excluded or minimized in controlled trials and are rarely captured in claims-based real-world studies, yet they are essential for understanding obesity care in practice. Third, it describes real-world program retention, medication coverage, and treatment patterns (dosing, escalation, and switching), and shows how a continuous remote care model can accommodate these trajectories while supporting weight loss through 12 months.

## Methods

### Study Setting and Objectives

This retrospective real-world cohort study evaluated the effectiveness of two medication-supported treatment (Rx) pathways within Virta’s comprehensive obesity care program offered through employer and health plan clients. Participants receive individualized medical management from licensed clinicians together with continuous remote support from health coaches.

### Data Source

Data were extracted from Virta Health’s program database and electronic health record, which contain clinician documentation, laboratory results, weight and biomarker data uploaded through the Virta mobile application (app) from bluetooth-connected devices, and other program-related information, including app-recorded side-effect data. The analysis used de-identified data.

### Program Setting and Organization

Eligible members enroll voluntarily and are matched to a product pathway based on clinical needs and client benefit design. Each participant receives individualized medical management from a licensed clinician (physician, nurse practitioner, or both) and continuous remote support from a health coach. Through the app, participants can message their care team, access educational content, and log biomarkers, side effects, and weight, the latter via a bluetooth-connected scale. For individuals with overweight or obesity, the program offers three different product pathways: VINT only, VINT+IOM, and IOM only. This study evaluated the two Rx pathways (VINT+IOM and IOM only), which are available only through clients whose benefit design covers IOMs.

### Treatment Pathways

The two pathways shared the same telemedicine infrastructure, including clinician-guided medication management, health coach access, GLP-1 education (prescribing process and side-effect management), and physical activity and strength-training content. They differed in the content and individualization of nutrition support.

Participants in the VINT+IOM pathway received an individualized carbohydrate-reduced nutritional intervention delivered through Virta’s remote care model. The intervention emphasized reducing refined carbohydrates and added sugars while prioritizing protein-rich foods, nonstarchy vegetables, and dietary fats to support satiety and nutritional adequacy. Carbohydrate recommendations were individualized according to participants’ clinical characteristics, metabolic health, treatment goals, and tolerability, rather than prescribing a uniform macronutrient distribution. Participants had access to meal-planning and grocery tools and monitored blood beta-hydroxybutyrate (BHB) as an objective marker of carbohydrate reduction; however, no specific calorie target was prescribed. Health coaches supported dietary implementation and adherence, while clinicians monitored progress and adjusted medications as clinically indicated.

Participants in the IOM-only pathway received care through the same infrastructure, including clinician oversight, medication management, and coach access. Their nutrition support consisted of general healthy-eating education covering protein, carbohydrate quality, added sugars, and dietary fats, together with behavioral guidance on habits, sleep, and stress. No carbohydrate targets, macronutrient distribution, calorie targets, or ketone monitoring were prescribed.

Dietary intake, and macronutrient distribution were not systematically collected in either pathway at the time the program was first offered, except for adherence to carbohydrate reduction through BHB logging in VINT+IOM. Food-logging features were introduced in a later rollout of the program and are not part of this current analytic cohort. Because both pathways operated within the same care system using similar medication management protocols, nutrition support was the principal difference between them.

### Pathway Assignment and Switching

Members were assigned to a pathway at enrollment based on clinical eligibility and participant choice, following shared decision-making with the care team. Pathway selection was not randomized. Members could change pathways at any time during follow-up, for example by switching between Rx pathways or moving between an Rx pathway and VINT-only care. We defined a pathway switch as a documented change in assigned product pathway, and every switch step is appropriately documented with a start and end date.

### Study Population

Eligible members were adults (aged ≥18 years) who enrolled in the program between October 1, 2024, and July 1, 2025, through employer or health plan clients whose benefit design included access to the Rx pathways. To ensure complete outcome ascertainment, the cohort was restricted to individuals who were expected to reach 12 months of follow-up by the data extraction date (July 1, 2026) and who were retained in the program through month 12.

Each members’ primary pathway was defined as their longest continuous enrollment episode in a product pathway (see Pathway Assignment and Switching), measured from the episode start date to its end date or, for ongoing enrollment, to the data extraction date. Retention through month 12 was determined at the program level, and participants were assigned to the primary Rx pathway of their longest episode regardless of its duration. Participants were classified as Rx VINT+IOM or IOM only pathways according to this primary pathway. The final analytic cohort was then restricted to participants who received at least one IOM prescription and whose longest episode in a primary Rx pathway lasted at least 9 months (≥270 days). Participants whose longest episode was in the nutrition-only pathway (Figure 1, step 2), or whose longest Rx episode was shorter than 9 months, were excluded (Figure 1, step 5).

**Figure 1.**
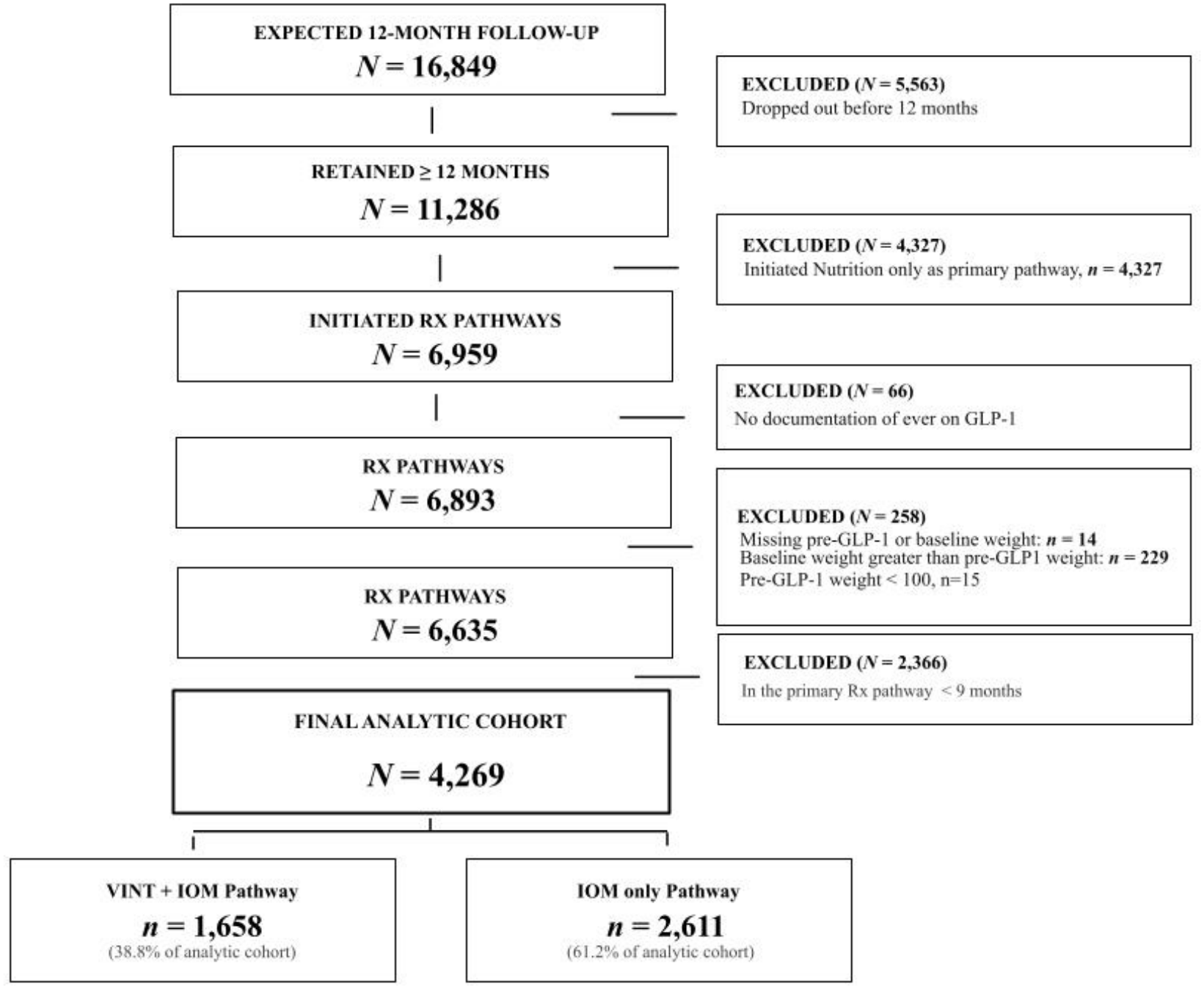
Flow diagram of Patient Selection Flow chart summarizing longitudinal patient retention, exclusion criteria, and final primary intervention pathway allocation. Abbreviations: GLP-1, glucagon-like peptide-1 receptor agonist; Rx, prescription pathway. Maintenance of assigned primary Rx pathway for a minimum of 9 months was required for inclusion in the final analytic cohort.

For participants already receiving an IOM at enrollment, the pre-GLP-1 weight was the self-reported weight, recalled at enrollment and documented in clinician notes, from before GLP-1 initiation. Participants with implausible pre-GLP-1 weights were excluded, defined as a pre-GLP-1 weight below 100 lb or, among participants already receiving an IOM at enrollment, a pre-GLP-1 weight lower than the enrollment (baseline) weight. Cohort derivation is shown in Figure 1.

### Measures and Variable Definitions

#### Index date and follow-up

Baseline was the program enrollment date, and baseline weight was the enrollment weight in the program database. Follow-up weights at 6 and 12 months were the average of weights recorded within ±30 days of day 180 and day 365 of the participant’s primary Rx pathway episode, respectively.

#### GLP-1 status

Participants were classified as having prior GLP-1 exposure if they were receiving a GLP-1 receptor agonist at program enrollment and had a documented pre-GLP-1 weight in clinician notes. All other participants were classified as GLP-1–naïve, that is, participants not receiving a GLP-1 at enrollment

#### Medication data

Medication data were extracted from the structured medication records in the electronic health record. We identified records for semaglutide, tirzepatide, and liraglutide, and extracted the ingredient, dose start and end dates, route, and daily dose. For each participant, we derived the earliest and latest dose dates, the ingredients used, and the number of distinct ingredients. Dose escalation was assessed monthly: for each ingredient, the highest daily dose recorded in a calendar month was compared with that of the previous month with a recorded dose, and a month was classified as an escalation month if the dose was higher. We summarized the number and percentage of escalation months, the maximum dose reached, and the time from the first dose to the maximum dose. Medication coverage was assessed using the proportion of days covered (PDC), calculated as the number of unique days covered by a GLP-1 dose record divided by the number of days in the 12-month period after the start of the primary Rx pathway. Overlapping records and ingredient switches were counted once per day, and PDC≥80% was considered high coverage.

#### Side effects

Participants self-reported IOM–related side effects through the app as part of daily symptom and biomarker logging. Reported symptoms included fatigue, nausea, vomiting, diarrhea, constipation, and abdominal pain. Each entry was date-stamped, and we extracted the daily symptom values and the number of entries per symptom. Side effects were summarized descriptively. Because logging was voluntary and participant-initiated, the data reflect reported symptoms rather than clinician-adjudicated adverse events.

### Outcomes

#### Primary outcomes

The primary outcome was 12-month weight change from program enrollment, evaluated using both adjusted body weight change derived from longitudinal mixed-effects models and observed percentage weight loss. Clinically meaningful weight-loss response was assessed as the proportion of participants achieving ≥5%, ≥10%, ≥15%, and ≥20% weight loss at 12 months.

The primary analysis focused on GLP-1–naïve participants who received a single GLP-1–based active pharmaceutical ingredient (semaglutide or tirzepatide) throughout the 12-month intervention period to minimize the potential influence of prior medication exposure and ingredient switching on weight-loss outcomes. Ancillary analyses evaluated weight-loss outcomes in the overall prescription cohort and across additional subgroups, including the broader GLP-1–naïve population, participants with prior GLP-1 exposure, and those who switched medication ingredients during follow-up. Among participants with prior GLP-1 exposure, weight-loss outcomes were further examined separately for single active pharmaceutical ingredient semaglutide and tirzepatide users. Cumulative weight loss from the pre-GLP-1 reference weight through program enrollment and 12-month follow-up was also evaluated to characterize weight-loss trajectories before and after enrollment.

#### Secondary outcomes

Secondary outcomes included real-world medication utilization and management (medication adherence, medication ingredient, dose titration, ingredient switching, and documented medication trajectory), a descriptive assessment of reported medication side effects, and exploratory analyses of weight-loss outcomes according to medication adherence assessed by PDCs and ingredient switching.

#### Exploratory outcome

An exploratory outcome was change in HbA1c from program enrollment to 6 and 12 months among participants in the primary single-ingredient, GLP-1–naïve cohort with available HbA1c values.

#### Statistical Analysis

Baseline characteristics were summarized using descriptive statistics. Continuous variables are presented as means ± standard deviations (SD) or medians with interquartile ranges (IQR), as appropriate, whereas categorical variables are presented as counts and percentages. Baseline characteristics were compared between treatment pathways using Student’s *t* test or the Wilcoxon rank-sum test for continuous variables and the chi-square test or Fisher’s exact test for categorical variables, as appropriate.

Longitudinal linear mixed-effects models were used to estimate adjusted body weight trajectories and estimated marginal means (EMMs) with 95% confidence intervals at program enrollment, 6 months, and 12 months. Models included time, treatment pathway, and their interaction, with adjustment for baseline age, sex, BMI, GLP-1 medication use, and HbA1c to account for differences in enrollment characteristics between treatment pathways. Participant-level random intercepts were included to account for repeated measurements within individuals. Adjusted weight change from program enrollment to 12 months was estimated for each treatment pathway and prespecified subgroup, allowing comparisons of weight-loss outcomes while accounting for measured baseline differences between groups. To examine sex differences, we extended the adjusted linear mixed effect models (random participant intercept; adjusted for age, baseline HbA1c and baseline BMI) with time × sex, time × sex × pathway, time × sex × medication, and time × sex × pathway × medication terms. Sex-specific adjusted means and changes from program start were estimated with model-based marginal means, and differences in change between women and men were tested within each medication–pathway group. Analyses included women and men only.

HbA1c was analyzed as an exploratory outcome in the primary single-ingredient, GLP-1–naïve cohort, restricted to participants with at least one available HbA1c value. HbA1c was modeled with a linear mixed-effects model including time (enrollment, 6 months, 12 months), treatment pathway, medication (semaglutide or tirzepatide), and their three-way interaction, with adjustment for baseline age, sex, and BMI. Baseline HbA1c was not included as a covariate because it was the first observation of the outcome. Participant-level random intercepts accounted for repeated measurements.

Observed absolute weight change, percentage weight loss, and average monthly weight loss (calculated descriptively as adjusted 12-month weight change divided by 12 months) were summarized using available body weight measurements within each follow-up window. Between-group comparisons of observed weight outcomes were performed using the Wilcoxon rank-sum test. Weight-loss responder analyses evaluated the proportions of participants achieving ≥5%, ≥10%, ≥15%, and ≥20% weight loss at 12 months. Differences in responder proportions between pathways were compared using the chi-square test or Fisher’s exact test, as appropriate.

Medication utilization and management were summarized descriptively and included starting and final medication ingredients and doses, dose escalation and de-escalation, ingredient switching, medication trajectories, documented medication continuation, proportion of days covered (PDC), and reported medication side effects. Continuous medication-related variables were compared using the Wilcoxon rank-sum test, whereas categorical medication utilization variables were compared using the chi-square test or Fisher’s exact test, as appropriate. All statistical analyses were performed using R (version 4.6.1; R Foundation for Statistical Computing, Vienna, Austria). Statistical significance was defined as a two-sided P < 0.05.

## Results

### Study population and enrollment characteristics

Among participants expected to have 12 months of follow-up, 11,286 were retained through 12 months (Figure 1). Retention was high across both Rx pathways, with 77% (2,642/3,426) of participants in VINT+IOM and 72% (4,317/6,002) of those in IOM-only retained through 12 months. Following application of the study eligibility criteria and data-quality exclusions, 4,269 participants comprised the final analytic cohort, including 1,658 (38.8%) in VINT+IOM and 2,611 (61.2%) in IOM only (Figure 1).

Enrollment characteristics of the analytic cohort are presented in Table 1. Participants had a mean age of 47.7 years, 76.2% were female, and the median baseline BMI was 34.3 kg/m². Approximately one-half (52.6%) were receiving GLP-1 therapy at program entry. Compared with participants in the IOM-only pathway, those in the VINT+IOM pathway had higher baseline body weight (220 vs. 209 lb; SMD=0.20), higher BMI (35.6 vs. 33.6 kg/m²; SMD=0.26), were less likely to be receiving GLP-1 therapy at baseline (46.0% vs. 56.8%; SMD=0.22) and had slightly higher baseline HbA1c (5.5% vs. 5.4%; SMD=0.20). To account for these observed differences, longitudinal weight-loss analyses were adjusted for baseline demographic and clinical characteristics, including age, sex, BMI, GLP-1 medication use, and HbA1c. Most other demographic and laboratory characteristics were generally comparable between pathways. Overall, mean baseline HbA1c (5.4%), HDL-C (53.3 mg/dL), total cholesterol (186.7 mg/dL), ALT (24.1 U/L), AST (21.4 U/L), and ALP (75.7 U/L), and median triglycerides (102 mg/dL), were within the reference ranges (Table 1 footnote), whereas mean LDL-C (111.3 mg/dL) was slightly above the optimal threshold of 100 mg/dL.

**Table 1.** Baseline Characteristics of the Analytic Cohort by SWL Prescription Pathway.

| Characteristic | Overall<br>(N=4,269) | IOM only<br>(n=2,611) | VINT+IOM<br>(n=1,658) | P value | SMD |
|---|---|---|---|---|---|
| Demographic characteristics |  |  |  |  |  |
| Age, years | 47.7 (10.6) | 47.4 (10.6) | 48.0 (10.5) | 0.070 | 0.057 |
| Gender, n (%) |  |  |  | 0.683 | 0.029 |
| Female | 3251 (76.2) | 1986 (76.1) | 1265 (76.3) |  |  |
| Male | 1013 (23.7) | 621 (23.8) | 392 (23.6) |  |  |
| Other | 5 (0.1) | 4 (0.2) | 1 (0.1) |  |  |
| Non-Hispanic ethnicity, n (%) | 3815 (89.4) | 2365 (90.6) | 1450 (87.5) | 0.001 | 0.100 |
| Race, n (%) |  |  |  |  | **0.122** |
| American Indian or Alaska Native | 1 (0) | 0 (0) | 1 (0.1) |  |  |
| Asian | 77 (1.8) | 48 (1.8) | 29 (1.7) |  |  |
| Black | 637 (14.9) | 351 (13.4) | 286 (17.2) |  |  |
| Other or multiple races | 69 (1.6) | 42 (1.6) | 27 (1.6) |  |  |
| Other or unknown race | 218 (5.1) | 125 (4.8) | 93 (5.6) |  |  |
| White | 3267 (76.5) | 2045 (78.3) | 1222 (73.7) |  |  |
| Anthropometric and medication characteristics |  |  |  |  |  |
| Baseline weight, kg | 96.6 [82.1, 113.4] | 94.8 [80.7, 112.0] | 99.8 [85.3, 115.7] | <0.001 | 0.196 |
| Baseline weight, lb | 213 [181, 250] | 209 [178, 247] | 220 [188, 255] | <0.001 | 0.196 |
| Baseline BMI, kg/m² | 34.3 [30.1, 39.1] | 33.6 [29.6, 38.4] | 35.6 [31.2, 40.3] | <0.001 | 0.260 |
| Receiving GLP-1 therapy at baseline, n (%) | 2245 (52.6) | 1482 (56.8) | 763 (46) | <0.001 | 0.216 |
| Missing pre-GLP-1 weight, n (%) | 3 (0.1) | 2 (0.1) | 1 (0.1) | 1.000 | 0.006 |
| Pre-GLP-1 weight, kg | 105.7 [92.5, 122.5] | 105.2 [92.1, 122.5] | 106.6 [93.0, 122.5] | 0.260 | 0.027 |
| Pre-GLP-1 weight, lb | 233 [204, 270] | 232 [203, 270] | 235 [205, 270] | 0.260 | 0.027 |
| Number of GLP-1 medication types | 1.4 (0.5) | 1.4 (0.5) | 1.4 (0.5) | 0.255 | 0.036 |
| Baseline complexity score | 1 [1, 3] | 3 [1, 3] | 1 [1, 3] | <0.001 | 0.284 |
| Program participation characteristics |  |  |  |  |  |
| Days in longest SWL pathway | 383 [335, 438] | 378 [330, 435] | 390 [344, 444] | <0.001 | 0.136 |
| Number of pathway switches | 2 [1, 3] | 2 [1, 3] | 2 [2, 3] | <0.001 | 0.279 |
| Remained in SWL pathway ≥270 days, n (%) | 4269 (100) | 2611 (100) | 1658 (100) | 1.000 | <0.001 |
| Remained in SWL pathway ≥365 days, n (%) | 2535 (59.4) | 1448 (55.5) | 1087 (65.6) | <0.001 | 0.208 |
| Baseline laboratory measures |  |  |  |  |  |
| HbA1c, % | 5.4 (0.4) | 5.4 (0.4) | 5.5 (0.5) | <0.001 | 0.198 |
| HDL cholesterol, mg/dL | 53.3 (15.3) | 53.2 (15.4) | 53.3 (15.3) | 0.936 | 0.003 |
| LDL cholesterol, mg/dL | 111.3 (32.9) | 110.4 (32.9) | 112.7 (32.9) | 0.031 | 0.072 |
| Total cholesterol, mg/dL | 186.7 (38.4) | 185.9 (38.6) | 187.8 (38.1) | 0.142 | 0.049 |
| Triglycerides, mg/dL | 102 [75, 142] | 102 [75, 144] | 103 [75, 140] | 0.946 | 0.012 |
| ALT, U/L | 24.1 (17.8) | 23.7 (17.0) | 24.7 (19.0) | 0.065 | 0.059 |
| AST, U/L | 21.4 (11.9) | 21.2 (11.2) | 21.7 (12.9) | 0.238 | 0.038 |
| Alkaline phosphatase, U/L | 75.7 (24.7) | 75.0 (24.3) | 76.8 (25.4) | 0.020 | 0.075 |

Data are presented as mean (SD), median [IQR], or n (%), unless otherwise indicated. P values compare IOM only and VINT+IOM. SMD indicates standardized mean difference; values of approximately 0.10, 0.20, and 0.50 are commonly interpreted as small, moderate, and large differences, respectively. Reference ranges: HbA1c, <5.7% normal, 5.7-6.4% prediabetes, ≥6.5% diabetes range; HDL-C, ≥40 mg/dL (men) and ≥50 mg/dL (women); LDL-C, <100 mg/dL; total cholesterol, <200 mg/dL; triglycerides, <150 mg/dL; ALT, [7-55 U/L]; AST, [8-48 U/L]; ALP, [44-147 U/L]. [Liver enzyme ranges are those of the performing] ALP, alkaline phosphatase; ALT, alanine aminotransferase; AST, aspartate aminotransferase; BMI, body mass index; GLP-1, glucagon-like peptide-1; HbA1c, hemoglobin A1c; HDL-C, high-density lipoprotein cholesterol; IOM, incretin-based obesity medication; IQR, interquartile range; LDL-C, low-density lipoprotein cholesterol; SD, standard deviation; SWL, Sustainable Weight Loss; VINT, Virta Individualized Nutrition Therapy.

### Primary Outcomes

#### Adjusted and Observed Weight-Loss Outcomes and Responder Rates

Weight loss was clinically meaningful in every cohort, and within-group reductions from program start to 12 months were significant in all groups (P<0.001; Table 2, Supplementary Table S1). In both adjusted (Table 2, Supplementary Table S1) and observed (Table 3) analyses, weight loss was consistently greater with VINT+IOM than with IOM only.

**Table 2.** Adjusted Estimated Marginal Mean Weight Outcomes Across Cohorts and Medication Subgroups.

| Pathway | Medication | Pre-GLP-1 EMM, kg (95% CI) | Program-start EMM, kg (95% CI) | 6-mo EMM, kg (95% CI) | 6-mo change, kg (95% CI) | 12-mo EMM, kg (95% CI) | 12-mo change, kg (95% CI) | WG P value | BG P value |
|---|---|---|---|---|---|---|---|---|---|
| **Primary outcomes** |  |  |  |  |  |  |  |  |  |
| **Single ingredient (GLP-1 naive)** |  |  |  |  |  |  |  |  |  |
| IOM | Semaglutide | — | 107.8 (101.3 to 114.3) | 101.2 (94.7 to 107.7) | −6.6 (−7.5 to −5.8) | 96.9 (90.4 to 103.3) | −11.0 (−11.8 to −10.1) | <0.001 | ref |
| IOM | Tirzepatide | — | 108.8 (102.1 to 115.4) | 100.0 (93.3 to 106.6) | −8.8 (−10.0 to −7.5) | 93.2 (86.5 to 99.8) | −15.6 (−16.9 to −14.4) | <0.001 | ref |
| VINT+IOM | Semaglutide | — | 108.1 (101.6 to 114.7) | 98.7 (92.1 to 105.2) | −9.4 (−10.4 to −8.5) | 94.2 (87.6 to 100.7) | −13.9* (−14.9 to −13.0) | <0.001 | 0.012 |
| VINT+IOM | Tirzepatide | — | 109.7 (103.0 to 116.4) | 97.3 (90.5 to 103.9) | −12.5 (−13.8 to −11.1) | 88.9 (82.2 to 95.6) | −20.8* (−22.2 to −19.5) | <0.001 | 0.003 |
| **Ancillary outcomes** |  |  |  |  |  |  |  |  |  |
| **Overall Rx cohort** |  |  |  |  |  |  |  |  |  |
| IOM | All | — | 102.6 (99.7 to 105.5) | 96.1 (93.3 to 99.0) | −6.5 (−6.8 to −6.1) | 92.9 (90.0 to 95.7) | −9.7 (−10.1 to −9.4) | <0.001 | ref |
| VINT+IOM | All | — | 102.6 (99.7 to 105.5) | 93.5 (90.7 to 96.4) | −9.1 (−9.5 to −8.7) | 89.3 (86.5 to 92.2) | −13.3* (−13.8 to −12.9) | <0.001 | <0.001 |
| **GLP-1 naive** |  |  |  |  |  |  |  |  |  |
| IOM | All | — | 104.2 (99.5 to 108.9) | 96.5 (91.9 to 101.2) | −7.6 (−8.2 to −7.1) | 91.8 (87.1 to 96.5) | −12.4 (−12.9 to −11.8) | <0.001 | ref |
| VINT+IOM | All | — | 104.1 (99.3 to 108.8) | 93.4 (88.7 to 98.2) | −10.6 (−11.3 to −10.0) | 88.2 (83.4 to 92.9) | −15.9* (−16.5 to −15.3) | <0.001 | <0.001 |
| **Prior GLP-1** |  |  |  |  |  |  |  |  |  |
| IOM | All | 116.0 (112.3 to 119.7) | 101.2 (97.5 to 104.8) | 95.6 (91.9 to 99.3) | −5.5 (−6.1 to −5.0) | 93.5 (89.8 to 97.2) | −7.7 (−8.3 to −7.1) | <0.001 | ref |
| VINT+IOM | All | 112.9 (109.2 to 116.6) | 101.2 (97.5 to 105.0) | 94.0 (90.3 to 97.7) | −7.3 (−8.1 to −6.5) | 90.9 (87.2 to 94.7) | −10.3* (−11.1 to −9.5) | <0.001 | <0.001 |
| **Single ingredient (Prior GLP-1)** |  |  |  |  |  |  |  |  |  |
| IOM | Semaglutide | 115.2 (111.5 to 118.9) | 99.8 (96.1 to 103.5) | 95.3 (91.6 to 99.0) | −4.5 (−5.4 to −3.7) | 93.8 (90.1 to 97.5) | −6.0 (−6.8 to −5.1) | <0.001 | ref |
| IOM | Tirzepatide | 114.8 (110.8 to 118.8) | 100.2 (96.3 to 104.2) | 92.0 (88.0 to 96.0) | −8.2 (−9.6 to −6.8) | 88.1 (84.1 to 92.1) | −12.1 (−13.5 to −10.7) | <0.001 | ref |
| VINT+IOM | Semaglutide | 111.8 (108.0 to 115.7) | 99.7 (95.9 to 103.6) | 93.3 (89.5 to 97.2) | −6.4 (−7.5 to −5.3) | 90.6 (86.8 to 94.4) | −9.1* (−10.2 to −8.0) | <0.001 | 0.018 |
| VINT+IOM | Tirzepatide | 111.1 (106.8 to 115.4) | 99.9 (95.6 to 104.1) | 90.5 (86.2 to 94.8) | −9.4 (−11.3 to −7.5) | 85.4 (81.1 to 89.7) | −14.5* (−16.4 to −12.5) | <0.001 | 0.035 |

*Notes: Values were converted from pounds to kilograms (1 lb = 0.45359 kg) and rounded to one decimal. Values are adjusted estimated marginal means (EMMs) with 95% confidence intervals from longitudinal mixed-effects models. Negative changes indicate weight loss from program start; 95% CIs for the changes are from the model contrasts (program start vs 6 and 12 months). All program-start to 12-month within-group changes were statistically significant (P < 0.001). \* Indicates a statistically significant between-group difference (VINT+IOM vs. IOM only within the respective cohort/medication subgroup, P < 0.05). 'ref' indicates the baseline reference group for between-group P-value comparisons. Abbreviations: VINT, Virta Individualized Nutrition Intervention; IOM, Incretin based obesity medication; mo, month; WG, within-group; BG, between-group.*

**Table 3.** Observed Percentage Weight Loss and 12-Month Weight-Loss Responders.

| **Analysis population** | **Pathway** | **Medication** | **Cohort N** | **6-mo N** | **Mean % 6 mo** | **12-mo N** | **Mean % 12 mo** | **≥5% weight loss,<br>n (%)** | **≥10% weight loss,<br>n (%)** | **≥15% / ≥20% weight loss,<br>n (%)** |
|---|---|---|---|---|---|---|---|---|---|---|
| **Primary outcomes** |  |  |  |  |  |  |  |  |  |  |
| **Single ingredient (GLP-1 naive)** |  |  |  |  |  |  |  |  |  |  |
|  | IOM | Semaglutide | 430 | 424 | 6.9% | 427 | 11.1% | 318 (74.5%) | 242 (56.7%) | 149 (34.9%) / 69 (16.2%) |
|  | IOM | Tirzepatide | 223 | 222 | 8.9% | 222 | 15.6% | 186 (83.8%) | 159 (71.6%) | 123 (55.4%) / 81 (36.5%) |
|  | VINT+IOM | Semaglutide | 360 | 359 | 9.2% | 356 | 13.3% | 309 (86.8%) | 230 (64.6%) | 142 (39.9%) / 70 (19.7%) |
|  | VINT+IOM | Tirzepatide | 202 | 201 | 12.1% | 200 | 19.7% | 193 (96.5%) | 169 (84.5%) | 135 (67.5%) / 91 (45.5%) |
| **Ancillary outcomes** |  |  |  |  |  |  |  |  |  |  |
| **Overall Rx cohort** |  |  |  |  |  |  |  |  |  |  |
|  | IOM | All | 2611 | 2578 | 6.5% | 2580 | 9.6% | 1,732 (67.1%) | 1,216 (47.1%) | 730 (28.3%) / 372 (14.4%) |
|  | VINT+IOM | All | 1658 | 1651 | 8.8% | 1646 | 12.7% | 1,291 (78.4%) | 965 (58.6%) | 643 (39.1%) / 375 (22.8%) |
| **GLP-1 naive** |  |  |  |  |  |  |  |  |  |  |
|  | IOM | All | 1129 | 1114 | 7.6% | 1116 | 12.1% | 858 (76.9%) | 657 (58.9%) | 424 (38.0%) / 230 (20.6%) |
|  | VINT+IOM | All | 895 | 891 | 10.3% | 885 | 15.2% | 783 (88.5%) | 623 (70.4%) | 442 (49.9%) / 261 (29.5%) |
| **Prior GLP-1** |  |  |  |  |  |  |  |  |  |  |
|  | IOM | All | 1482 | 1464 | 5.6% | 1464 | 7.7% | 874 (59.7%) | 559 (38.2%) | 306 (20.9%) / 142 (9.7%) |
|  | VINT+IOM | All | 763 | 760 | 7.1% | 761 | 9.8% | 508 (66.8%) | 342 (44.9%) | 201 (26.4%) / 114 (15.0%) |
| **Single ingredient (Prior GLP-1)** |  |  |  |  |  |  |  |  |  |  |
|  | IOM | Semaglutide | 677 | 668 | 4.8% | 669 | 6.3% | 357 (53.4%) | 215 (32.1%) | 105 (15.7%) / 40 (6.0%) |
|  | IOM | Tirzepatide | 240 | 240 | 8.6% | 240 | 12.5% | 190 (79.2%) | 145 (60.4%) | 91 (37.9%) / 53 (22.1%) |
|  | VINT+IOM | Semaglutide | 349 | 348 | 6.5% | 349 | 8.8% | 225 (64.5%) | 146 (41.8%) | 81 (23.2%) / 39 (11.2%) |
|  | VINT+IOM | Tirzepatide | 120 | 118 | 9.2% | 119 | 13.8% | 95 (79.8%) | 74 (62.2%) | 47 (39.5%) / 35 (29.4%) |

*Note. Percentage weight loss and responder outcomes are observed values among participants with an available weight in the specified follow-up window. For the prior GLP-1 cohort, outcomes are calculated from program start; total weight loss from the pre-GLP-1 reference is reported separately. The final column reports ≥15% and ≥20% responders, respectively.*

In the primary analysis of GLP-1–naïve participants receiving a single ingredient, VINT+IOM was associated with significantly greater adjusted 12-month weight loss than IOM only with both semaglutide (additional 3.0 kg [6.6 lb], 95% CI 0.6 to 5.4 kg [1.4 to 11.8 lb]; 1.2 vs 0.9 kg/month [2.6 vs 2.0 lb/month]; P=0.012) and tirzepatide (additional 5.2 kg [11.5 lb], 95% CI 1.8 to 8.7 kg [3.9 to 19.1 lb]; 1.7 vs 1.3 kg/month; P=0.003) (Figure 2A–B). Observed mean weight loss was 13.3% vs 11.1% with semaglutide and 19.7% vs 15.6% with tirzepatide. Responder rates were higher with VINT+IOM at every threshold (Table 3; Figure 2C–D). Weight continued to decline between 6 and 12 months in both pathways, with no plateau (Supplementary Figure S1). From approximately 6 to 12 months, adjusted weight trajectories remained downward, with estimated slopes ranging from −0.73 to −0.76 kg/month for semaglutide and −1.14 to −1.38 kg/month for tirzepatide (all P<0.001), indicating continued weight reduction during the latter half of follow-up.

**Figure 2.**
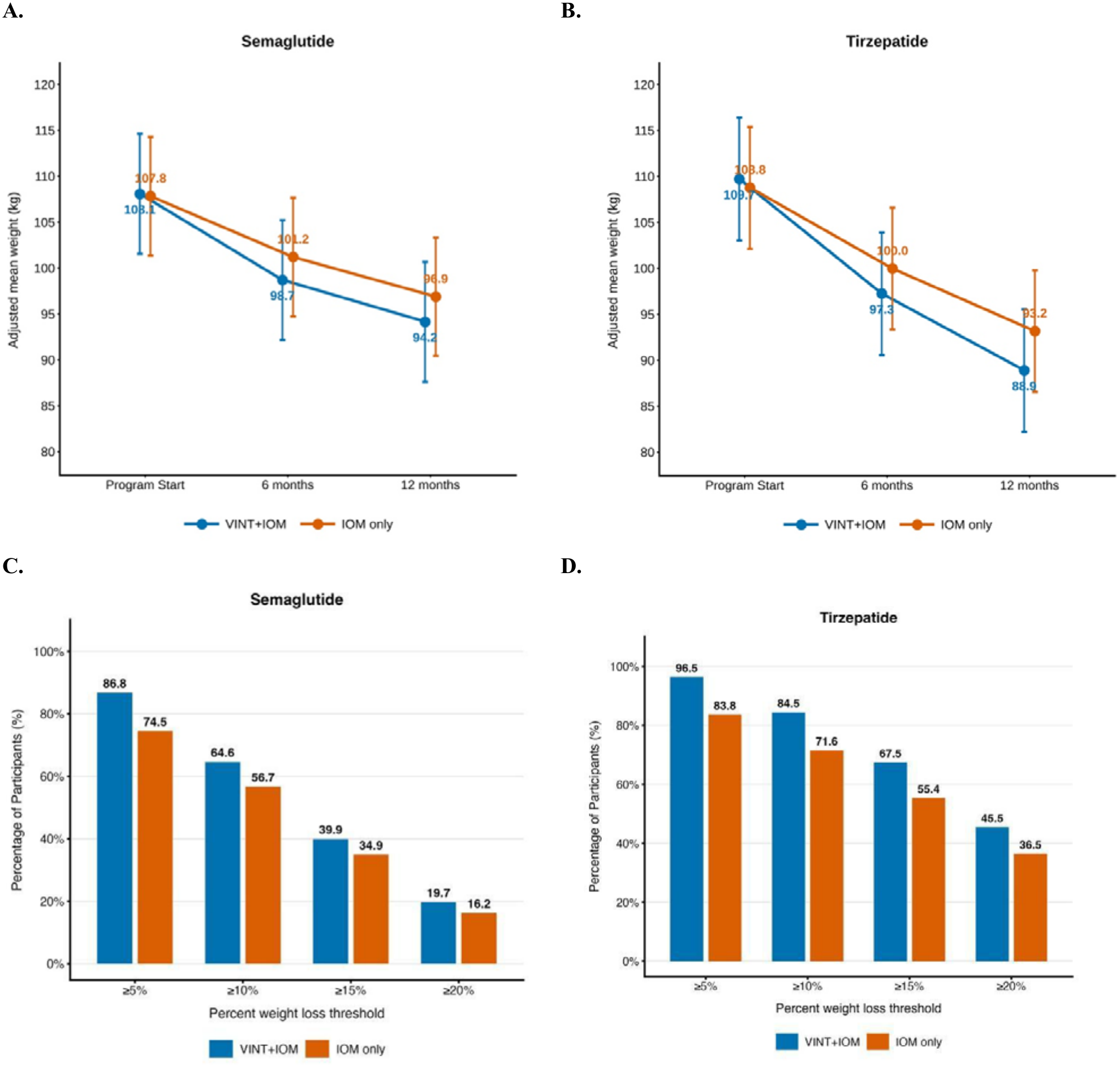
Adjusted Weight Trajectories and Weight-Loss Responder Rates at 12 Months Among GLP-1–Naïve, Single Active Ingredient Semaglutide and Tirzepatide Users by Intervention Pathway. **(A–B) Adjusted mean weight trajectories over 12 months.** Estimated marginal means (EMMs) of body weight (kg) at program start, 6 months, and 12 months for participants receiving (A) semaglutide or (B) tirzepatide, stratified by intervention pathway (VINT+IOM [blue lines] vs IOM only [orange lines]). Estimates were derived from linear mixed-effects models adjusted for baseline weight and demographic covariates. Points represent adjusted mean body weight, error bars indicate 95% confidence intervals (CIs), and point annotations display adjusted mean weight (kg) at each time point. **(C–D) Weight-loss responder rates at 12 months.** Proportions of participants achieving ≥5%, ≥10%, ≥15%, and ≥20% weight loss from program enrollment to 12 months among (C) semaglutide and (D) tirzepatide users, stratified by intervention pathway (VINT+IOM [blue bars] vs IOM only [orange bars]). Bars represent unadjusted percentages, with values above each bar indicating the proportion of participants achieving the corresponding weight-loss threshold.

#### Sex-Stratified Weight-Loss Outcomes and Sex-by-Treatment Interactions

Women made up 75.1% of the primary single-ingredient, GLP-1–naïve cohort (912 of 1,214). In adjusted mixed models including 683 women and 231 men, weight change over time differed by sex (time × sex, P=0.003; Supplementary Table S2), with men losing more absolute weight and starting 18–25 kg (39–54 lb) heavier (Supplementary Table S3; Supplementary Figure S2). The sex difference did not depend on medication (time × sex × medication, P=0.89), and the pathway interaction was not robust (P=0.039 alone, P=0.09 in the full model). Men’s additional loss was concentrated in VINT+IOM, where they lost 2.8 kg (6.1 lb) more than women at 12 months with semaglutide (95% CI 0.6 to 4.9 kg [1.3 to 10.9 lb]; P=0.012) and 3.0 kg (6.6 lb) more with tirzepatide (95% CI −0.1 to 6.1 kg [−0.2 to 13.4 lb]; P=0.059). In IOM only, sex differences were small and not significant (0.6 and 1.2 kg [1.3 and 2.6 lb]; P≥0.43; Supplementary Table S4).

In observed data, mean 12-month percentage weight loss was similar in women and men (14.3% vs 13.2%), as were the proportions achieving ≥5% (84.1% vs 82.1%), ≥10% (67.2% vs 64.1%) and ≥15% (47.1% vs 41.2%) weight loss (all P≥0.08; Supplementary Table S5). A higher proportion of women reached ≥20% weight loss (27.4% vs 21.3%; P=0.040). Within medication–pathway groups, this was significant only for semaglutide in IOM only (18.4% vs 9.5%; P=0.033)

#### Ancillary analyses

The advantage of VINT+IOM was maintained in the overall prescription cohort (additional 3.6 kg [7.9 lb], 95% CI 2.7 to 4.4 kg [6.0 to 9.8 lb]; 1.1 vs 0.8 kg/month [2.5 vs 1.8 lb/month]; observed loss 12.7% vs 9.6%; P<0.001; Figure 3) and among all GLP-1–naïve participants, including those who switched ingredients (additional 3.6 kg [7.9 lb], 95% CI 2.4 to 4.7 kg [5.4 to 10.4 lb]; 1.3 vs 1.0 kg/month [2.9 vs 2.3 lb/month]; 15.2% vs 12.1%; P<0.001; Table 2), with higher responder rates at all thresholds (Table 3). Among participants with prior GLP-1 exposure, both pathways achieved further weight loss during the program, again greater with VINT+IOM (additional 2.6 kg [5.8 lb], 95% CI 1.5 to 3.8 kg [3.3 to 8.3 lb]; 9.8% vs 7.7% from program start; P<0.001), including among single-ingredient semaglutide and tirzepatide users (Table 2). From the pre-GLP-1 reference weight, cumulative loss in VINT+IOM reached 19.7% overall at 12 months (Supplementary Table S6), 19.2% among semaglutide users, and 23.0% among tirzepatide users at 12 months. Correspondingly, 66.9% and 45.7% of VINT+IOM participants achieved ≥15% and ≥20% cumulative weight loss, respectively. Among semaglutide users, these proportions were 66.2% and 42.7%, while among tirzepatide users, 76.5% and 59.7% achieved these respective thresholds.

**Figure 3.**
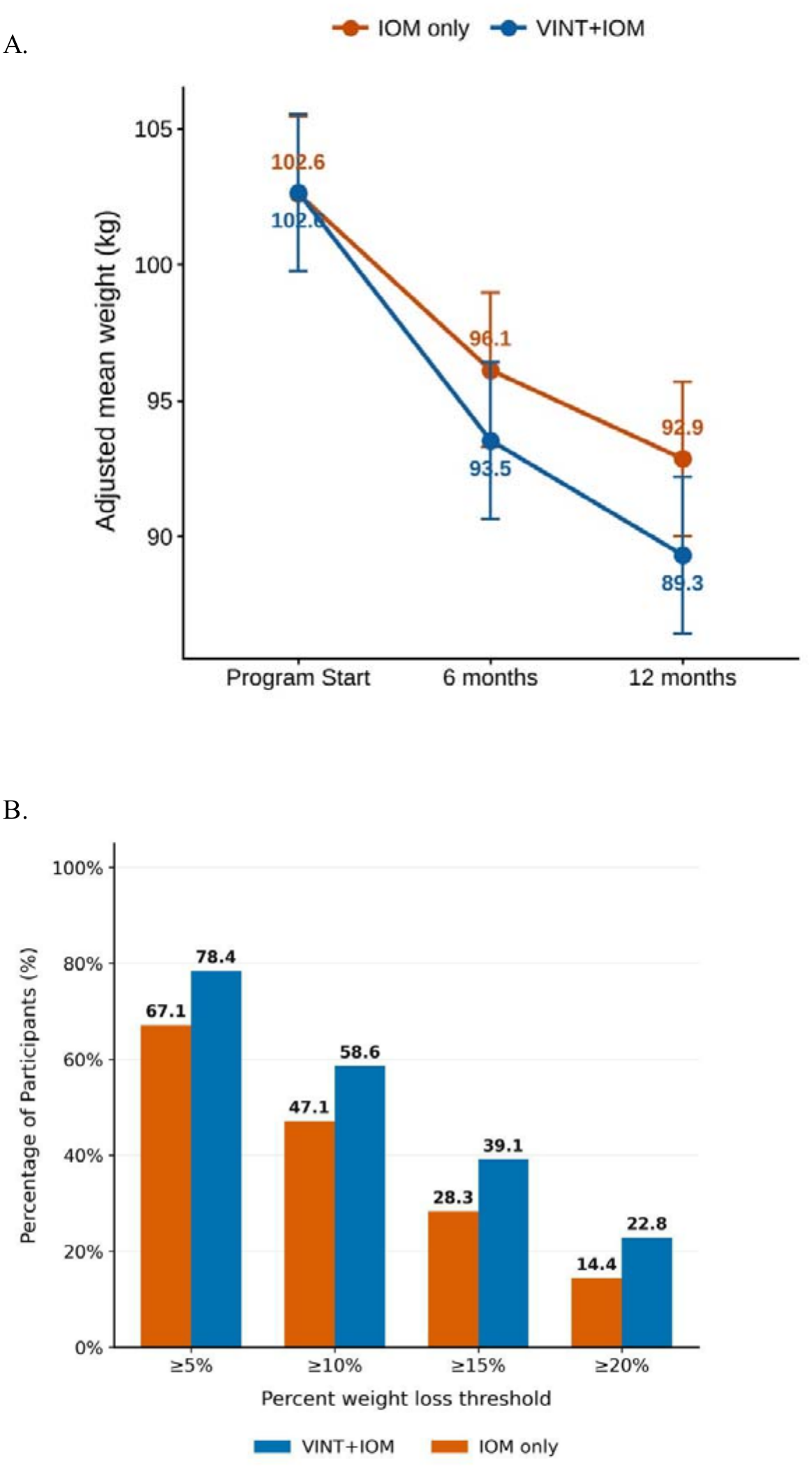
Adjusted Weight Trajectories and Weight-Loss Responder Rates at 12 Months in the Overall Prescription Cohort by Intervention Pathway. **(A) Adjusted mean weight trajectories over 12 months.** Estimated marginal means (EMMs) of body weight (kg) at program start, 6 months, and 12 months in the overall prescription cohort, stratified by intervention pathway (VINT+IOM [blue line] vs IOM only [orange line]). Estimates were derived from linear mixed-effects models adjusted for baseline weight and demographic covariates. Points represent adjusted mean body weight, error bars indicate 95% confidence intervals (CIs), and point annotations display adjusted mean weight (kg) at each time point. **(B) Weight-loss responder rates at 12 months.** Proportions of participants achieving ≥5%, ≥10%, ≥15%, and ≥20% weight loss from program enrollment to 12 months in the overall prescription cohort, stratified by intervention pathway (VINT+IOM [blue bars] vs IOM only [orange bars]). Bars represent unadjusted percentages, with values above each bar indicating the proportion of participants achieving the corresponding weight-loss threshold. *Abbreviations:* CI, confidence interval; EMM, estimated marginal mean; IOM, Incretin-based Obesity Medication; VINT, Virta Individualized Nutrition Therapy

### Medication exposure and management

The distribution of starting GLP-1 ingredients was similar between pathways, with approximately half of participants receiving semaglutide and half tirzepatide at the beginning of the observation period (Supplementary Table S7). However, among participants with prior GLP-1 exposure, those entering the IOM-only pathway generally initiated the program at higher medication doses than those entering VINT+IOM, suggesting they may have been further along in their GLP-1 treatment course before program enrollment.

Consistent with these lower starting doses, participants in VINT + IOM were more likely to undergo dose escalation during follow-up (84.8% vs 77.7%; *P*<0.001), experienced more dose increases, and required a longer median time to reach their maximum recorded dose (99 vs 76 days; both *P*<0.001) (Supplementary Tables S8-S10).

Medication coverage during follow-up was high in both pathways. In the overall cohort, mean PDC was 89.2% (SD, 16.0%) in VINT + IOM and 88.1% (SD, 16.5%) in IOM only, with median PDCs of 97.0% (IQR, 83.9%–100.0%) and 94.8% (IQR, 83.3%–99.5%), respectively. Although PDC distributions differed between pathways (*P*<0.001), the proportion of participants with PDC ≥80% was similar (80.5% vs 79.5%; *P*=0.44). Ingredient-specific analyses showed a higher PDC distribution with VINT + IOM among semaglutide users, whereas PDC distributions were similar between pathways among tirzepatide users. The proportion with PDC ≥80% did not differ significantly between pathways for either active ingredient (Supplementary Table S11). At 12 months, 80.0% of participants in VINT + IOM and 82.8% in IOM only pathway was documented as actively receiving GLP-1 therapy (*P*=0.02). Because medication exposure and status were derived from clinical medication records rather than pharmacy dispensing data, these findings should be interpreted as measures of documented medication coverage and management rather than pharmacy-dispensing adherence or persistence.

Medication regimens changed frequently over time, reflecting individualized clinical management. Approximately 38%-39% of participants switched GLP-1 ingredients during follow-up, with no significant difference between pathways. Among participants with a documented GLP-1 active ingredient switch, transitions occurred almost exclusively between semaglutide and tirzepatide (Supplementary Table S7). In IOM only, 789 of 1,028 participants with an ingredient switch (76.8%) transitioned from tirzepatide to semaglutide, whereas 230 (22.4%) transitioned from semaglutide to tirzepatide. Similarly, in VINT + IOM, 494 of 623 participants with an active ingredient switch (79.3%) transitioned from tirzepatide to semaglutide, and 121 (19.4%) transitioned from semaglutide to tirzepatide. Escalation-only trajectories were more common in VINT + IOM than in IOM only pathway (27.7% vs 20.9%), whereas ingredient switching occurred in 27.3% and 29.3% of participants, respectively (Supplementary Table S8). After validation of medication records, approximately 13% of participants in each pathway discontinued and subsequently restarted therapy, whereas confirmed discontinuation without a documented restart was uncommon (1.7% in VINT + IOM and 1.6% in Lifestyle + IOM). Active ingredient switching followed by confirmed discontinuation was similarly rare (3.2% vs 3.0%).

Despite differences in starting doses and titration patterns, medication intensity generally converged over follow-up. To further characterize dose exposure, starting and final doses were examined among GLP-1-naïve participants who received a single GLP-1 active ingredient during follow-up. Among semaglutide users, 66.1% of participants in VINT+IOM, and 55.8% in IOM only reached 2.4 mg as their final observed dose (*P*=0.003). Among tirzepatide users, 45.5% and 40.8% had 15 mg as their final observed dose (*P*=0.32) (Supplementary Table S10). Thus, a substantial proportion of participants in both pathways remained below the maximum maintenance doses commonly evaluated in obesity clinical trials, particularly for tirzepatide. Although VINT + IOM participants were somewhat more likely to reach and remain at the maximum semaglutide dose, differences in maximum-dose exposure were not consistently observed across medications.

### Weight loss across medication adherence and ingredient switching

In exploratory subgroup analyses, participants in the VINT+IOM pathway generally achieved greater 12-month weight loss than those in the IOM-only pathway across GLP-1 active ingredient-switching directions and medication adherence (PDC) categories (Supplementary Table S12). Among participants switching from tirzepatide to semaglutide, VINT+IOM was associated with significantly greater absolute (30.8 vs. 22.3 lb) and percentage weight loss (13.0% vs. 10.1%; both P<0.001). A similar pattern was observed among those switching from semaglutide to tirzepatide, with significantly greater percentage weight loss in the VINT+IOM pathway (8.5% vs. 6.5%; P=0.042). When stratified by medication adherence, participants in the VINT+IOM pathway also achieved significantly greater absolute weight loss in both the PDC <50% (27.8 vs. 20.2 lb; P=0.035) and PDC 50% to <80% categories (28.5 vs. 22.0 lb; P<0.001). Percentage weight loss was also significantly greater in the PDC 50% to <80% category (11.9% vs. 9.7%; P=0.004). Overall, these exploratory findings suggest a consistent pattern of greater weight loss with VINT+IOM across medication-switching and lower-adherence subgroups, although not all comparisons were statistically significant.

### Reported medication side effects

Reported medication side effects were similar between pathways (Supplementary Table S13). At least one side effect was documented for 64.6% of participants in VINT+IOM and 63.6% in IOM only (*P*=0.50). Among participants with any reported side effect, the median number of reporting days was 4 (IQR, 2–8), and the median number of distinct symptom types was 2 (IQR, 1–3). Approximately 74% of reported side effects occurred within the first 6 months after program enrollment. Nausea was the most frequently reported symptom in both pathways, followed by constipation, fatigue, diarrhea, and vomiting. With the exception of constipation, which was reported more frequently in VINT+IOM (37.2% vs. 32.8%; *P*=0.004), the frequencies of nausea, fatigue, diarrhea, vomiting, and other reported side effects were similar between pathways.

### Exploratory Outcome: HbA1c

In the primary single-ingredient, GLP-1–naïve cohort, HbA1c was available for 915 participants at enrollment, 395 at 6 months, and 479 at 12 months. Baseline HbA1c was in the normal range (about 5.4–5.5%). In adjusted mixed models, HbA1c decreased significantly in every medication and pathway group (all P<0.001; Supplementary Table S14), by 0.27 to 0.47 percentage points at 12 months. Reductions were greater with tirzepatide than with semaglutide (time × medication, P<0.001). Adjusted 12-month HbA1c did not differ significantly between VINT+IOM and IOM only, although the reduction was numerically larger with VINT+IOM for both medications. This small difference is expected given that baseline values were already in the normal range. Because follow-up HbA1c was available for a subset of members, these findings are exploratory.

## Discussion

In this real-world cohort of adults receiving incretin-based obesity medication within a continuous remote care program, adding individualized carbohydrate-reduced nutritional therapy to IOM (VINT+IOM) was consistently associated with greater adjusted 12-month weight loss than IOM alone. This held for GLP-1–naïve participants and those with prior GLP-1 exposure, and for both semaglutide and tirzepatide. Medication coverage and program retention were high in both pathways despite frequent dose changes and ingredient switching. These findings should be interpreted against established evidence that real-world obesity pharmacotherapy is considerably more dynamic than the standardized exposure achieved in randomized trials. Adherence, dose escalation, switching, and discontinuation vary substantially in practice, and this variability is frequently not captured in claims-based analyses. Earlier real-world studies reported mean PDCs of approximately 51%–54%, with only 27%–30% of participants achieving PDC ≥80% [1,2], and 12-month GLP-1 persistence estimates ranging from 24.5% in a large claims-based cohort to 58.6% for semaglutide and 64.8% for tirzepatide in more contemporary cohorts [1,2]. In contrast, approximately 80% of participants in our cohort achieved PDC ≥80%, and 12-month program retention was 77% with VINT+IOM and 72% with IOM only. Program retention and medication persistence are distinct measures and cannot be directly compared, but these results appear favorable. They are also consistent with an earlier analysis by Marshall et al. of commercially insured members who may have also received Virta-supported obesity care through the KeepWell program offered by Prime Therapeutics, in which 90.1% remained on GLP-1 therapy at 6 months with a mean weight reduction of 22.2 lb [20].

The additional weight loss with VINT+IOM (5.1–11.5 lb across cohorts, approximately 19%–52% greater than IOM only) is consistent with emerging, but still limited, evidence that nutritional strategies, particularly carbohydrate reduction, may add value alongside incretin therapy. HbA1c also declined in all groups, more with tirzepatide than with semaglutide and numerically more with VINT+IOM than with IOM only. However, differences between pathways were small and not statistically significant, which is unsurprising given that baseline values were normal (about 5.4-5.5%), leaving very little room for further reduction. In addition, limited follow-up laboratory data reduced the power to detect between-group differences. In a randomized pilot trial, Anyiam et al. found that semaglutide combined with a very-low-calorie diet produced approximately 7 percentage points greater weight loss than semaglutide alone, with greater fat-mass loss and improvements in metabolic measures [21]. Parker et al. similarly reported substantial weight loss with a personalized ketogenic diet plus low-dose semaglutide, with 92% of weight loss attributable to estimated fat mass and skeletal muscle mass largely preserved [22]. With tirzepatide, Schiavo et al. reported that a low-energy ketogenic intervention achieved similar overall weight loss but better preservation of fat-free mass, muscle strength, and resting metabolic rate than a conventional low-calorie diet [23], and a subsequent analysis also showed greater improvements in hepatic steatosis and stiffness [24]. A preliminary post hoc analysis of body composition data from a VINT clinical trial in individuals with T2D further suggests that a greater degree of carbohydrate reduction may support lean-mass preservation and physical function during weight loss [25]. Together, these studies raise the possibility that carbohydrate-reduced strategies may influence not only the magnitude of weight loss but also its composition, as well as other cardiometabolic outcomes, although most of the published studies on body composition were small or uncontrolled. Body composition was not assessed in our study, so we cannot determine whether the additional weight loss with VINT+IOM reflects fat or lean mass. Future studies that include body composition measures would help clarify whether carbohydrate-reduced nutritional therapy affects the quality, and not only the amount, of weight lost with incretin therapy.

Women made up three quarters of our cohort, so we examined whether sex modified these results. Randomized and real-world data generally show greater percentage weight loss with GLP-1 receptor agonists in women. A 2026 meta-analysis of sex-stratified trials found weight loss was greater among women (10.9%) than men (6.8%), and a real-world tirzepatide cohort reported greater weight loss in women than men at 15 months (15.1% vs 10.7%) [26,27]. We did not see this pattern. Men lost more absolute weight, which largely reflects their higher baseline weight, while observed 12-month percentage weight loss and most responder rates were similar by sex, and the sex difference did not vary by medication. A larger male advantage appeared only within VINT+IOM and was not consistent across models. One possible explanation is that men respond more strongly to the structured nutrition-therapy component, since a review of sex differences in obesity treatment reports that lifestyle modification, on average, produced larger weight losses in men [28]. We did not test this, and the finding should be treated as hypothesis-generating. Overall, the benefit of VINT+IOM over IOM alone was not restricted to one sex.

The magnitude of weight loss observed with VINT+IOM should be considered in the context of medication dose and duration of exposure. In pivotal efficacy trials, semaglutide was titrated by protocol toward the 2.4-mg maintenance dose, whereas in the present study initiation and titration occurred under routine clinical conditions. Among GLP-1–naïve participants who received semaglutide throughout follow-up, 66.1% had 2.4 mg as their final observed dose, and approximately one-third remained at lower doses. Observed weight loss in this group (13.3%) was similar in magnitude to the 14.9% reported at 68 weeks in STEP 1 [29] and to values reported in real-world studies of persistent semaglutide users maintained at 2.4 mg, including 14.1% at 12 months in SHAPE [30], 14.5% at 52 weeks in SCOPE [[31], and 17.6% at 12 months in WeGoTogether [32]. In addition, 12 months in VINT+IOM represents approximately 52 weeks of program participation rather than 52 weeks of semaglutide exposure, because participants-initiated treatment after enrollment and likely had shorter cumulative exposure, particularly to their final or higher doses.

A similar pattern was seen with tirzepatide. Among GLP-1–naïve participants who received tirzepatide throughout follow-up in VINT+IOM, 45.5% had 15 mg as their final observed dose, and cumulative exposure was likely less than 52 weeks. Weight loss of 19.7% was similar to the 19.5% and 20.9% reductions reported after 72 weeks with tirzepatide 10 and 15 mg, respectively, in SURMOUNT-1 [33], and higher than the 16.5% reported among tirzepatide users in SHAPE [30]. These observations indicate that substantial weight loss occurred in routine care among participants who did not reach maximum medication doses. They do not establish that the therapeutic effect is independent of medication dose or duration of exposure, and we did not evaluate dose-response relationships. In this setting, medication intensity was adjusted over time according to individual response and tolerability, alongside ongoing carbohydrate-reduced nutritional support from VINT. Among GLP-1–naïve participants, weight loss with VINT+IOM continued between 6 and 12 months without a clear plateau, suggesting that 12-month outcomes may not represent the maximal treatment response, particularly because initiation and titration occurred during program follow-up. However, these comparisons with clinical trials and other real-world studies should be interpreted cautiously. They are descriptive and involve differences in study populations, medication doses, treatment duration, follow-up, and analytic approach.

Substantial additional weight loss was also observed among participants who entered VINT + IOM with prior GLP-1 exposure. Among prior users remaining on a single ingredient, mean additional weight loss during 12 months of VINT + IOM was 8.8% with semaglutide and 13.8% with tirzepatide, despite pharmacotherapy and associated weight loss having begun before program entry. From the pre-GLP-1 reference, cumulative mean weight loss reached 19.2% and 23.0%, respectively, numerically exceeding the corresponding weight-loss estimates reported in pivotal clinical trials [26,30]. Notably, participants in the VINT+IOM pathway achieved approximately 12% weight loss at 12 months despite lower GLP-1 medication coverage (PDC <80%), with significantly greater absolute weight loss than IOM-only participants across both lower-PDC categories. This contrasts with real-world studies reporting attenuated weight loss with reduced GLP-1 exposure. A contemporary semaglutide study reported 6.4% weight loss among nonpersistent users and 7.2% among those following nonstandard titration patterns [34]. Similarly, another study reported 12-month weight loss of 4.7% with tirzepatide and 2.3% with semaglutide among early discontinuers, compared with 11.8% and 6.3%, respectively, among persistent users [35]. Although differences in populations, medication exposure, adherence definitions, and study designs preclude direct cross-study comparisons, the substantial weight loss observed with VINT+IOM despite prior GLP-1 exposure or inconsistent medication coverage suggests that carbohydrate-reduced nutritional therapy with longitudinal clinical support may remain clinically relevant across the variable medication trajectories encountered in routine obesity care. Given the observational design, however, the independent contribution of VINT cannot be established.

Taken together, these findings support conceptualizing IOM-based obesity treatment as a longitudinal and evolving care trajectory rather than a conventional prescription-centered approach characterized by intermittent clinical touchpoints and limited support between visits for navigating nutrition, adherence, persistence, tolerability, dose changes, switching, and treatment interruptions. In this study, both pathways included telemedicine infrastructure, clinician oversight, behavioral support, and access to IOM pharmacotherapy, while individualized carbohydrate-reduced nutrition therapy was the key additional component of VINT+IOM. The consistently greater weight loss observed with VINT+IOM, including among participants with prior GLP-1 exposure and across heterogeneous medication-use patterns, supports the potential incremental value of integrating individualized nutrition therapy within medication-supported obesity care. Importantly, 12-month program retention was high in both pathways (77% with VINT+IOM and 72% with IOM only), suggesting that structured carbohydrate-reduced nutritional therapy can be integrated with GLP-1–based pharmacotherapy while maintaining sustained participant engagement. These findings are consistent with previous real-world evidence demonstrating sustained engagement with VINT delivered independently of GLP-1–based pharmacotherapy (36).

These findings should be interpreted in the context of the observational study design. Treatment pathway selection was not randomized, and residual confounding, including differences in motivation, engagement, or other unmeasured characteristics, may have contributed to the observed between-group differences; therefore, the independent effect of VINT cannot be established. Although both pathways included health coach support, coaching frequency and intensity were not formally compared, and differences in coaching contact, particularly the dietary support provided in VINT+IOM, may have contributed to the observed differences. Medication selection, adherence, dose escalation, switching, and treatment duration were determined during routine clinical care rather than standardized by protocol, and analyses requiring continued pathway engagement and available weight measurements may have introduced selection bias toward more engaged participants. Because dose and duration of exposure were not randomized, the contribution of medication exposure cannot be separated from that of the nutrition pathway, and we did not evaluate dose-response relationships. Sex analyses were exploratory: they used absolute weight rather than percent change, included only 36–53 men per tirzepatide subgroup. We could not estimate effects for one participant with a third gender category, and we lacked data on menopausal or hormonal status, body composition, energy expenditure and dietary adherence, so we could not test the mechanisms that may underlie sex differences. Comparison with randomized trials and external real-world cohorts should therefore be considered contextual rather than direct. Finally, 12 months may be insufficient to characterize maximal weight loss or longer-term trajectories, particularly among participants initiating and titrating IOMs after enrollment. Longer follow-up is needed to determine weight-loss plateau and durability, medication continuation or discontinuation, and whether the differences observed between nutritional treatment pathways persist over time.

## Data Availability

All data produced in the present study are available upon reasonable request to the authors

## Supplementary Materials

The following supporting information can be downloaded from the journal website: Figure S1: Longitudinal body weight trajectories through 15 months among GLP-1–naïve participants receiving semaglutide or tirzepatide by treatment pathway; Figure S2: Adjusted mean body weight by sex over 12 months among GLP-1–naïve participants receiving a single GLP-1–based ingredient; Table S1: Adjusted estimated marginal mean weight outcomes across cohorts and medication subgroups; Table S2: Tests of sex-by-treatment interaction terms in the weight linear mixed models; Table S3: Observed and adjusted weight trajectories by sex, medication, and pathway; Table S4: Adjusted change in weight from program start by sex and sex difference in change; Table S5: Observed percentage weight loss and responder thresholds at 12 months by sex; Table S6: Total weight loss from the pre-GLP-1 reference among participants with prior GLP-1 exposure; Table S7: Medication utilization by Rx pathway; Table S8: Medication trajectories during 12 months; Table S9: Overall medication titration by Rx pathway; Table S10: Ingredient-specific medication doses by Rx pathway; Table S11: GLP-1 medication coverage by ingredient and Rx pathway among participants who stayed on a single ingredient; Table S12: Observed 12-month weight loss by GLP-1 ingredient-switching direction and proportion of days covered (PDC) category by Rx pathway; Table S13: Reported medication side effects during 12 months by treatment pathway; Table S14: Adjusted estimated marginal mean HbA1c outcomes by primary Rx pathway cohort and medication subgroup.

## Author Contributions

Conceptualization, S.J.A., and B.A.G.; methodology, S.J.A.; software, S.J.A.; validation, S.J.A., and A.R.Z.; formal analysis, S.J.A.; investigation, S.J.A.; resources, S.J.A.; data curation, S.J.A.; writing—original draft preparation, S.J.A.; writing—review and editing, S.J.A., A.R.Z., K.E.B., A.M.G., and B.A.G.; visualization, S.J.A.; supervision, B.A.G.; project administration, S.J.A. All authors have read and agreed to the published version of the manuscript.

## Funding

This research received no external funding.

## Institutional Review Board Statement

This retrospective observational study involved secondary analysis of an existing, de-identified clinical database. The database was determined to be exempt from IRB oversight by Advarra IRB (Pro00039234; 45 CFR 46.104(b)(4)). All analyses were conducted using de-identified patient information in accordance with the Health Insurance Portability and Accountability Act (HIPAA). No direct participant contact, research intervention, or additional data collection was undertaken. No institutional review board review or formal exemption determination was obtained.

## Informed Consent Statement

This study involved secondary analysis of existing, de-identified clinical data collected during routine clinical care. No study-specific informed consent was obtained, and no identifiable patient information was accessed for research purposes.

## Data Availability Statement

The data supporting the findings of this study were derived from de-identified clinical records collected during routine care within the Virta Health program. The individual-level datasets are not publicly available due to patient privacy, confidentiality, and proprietary data restrictions. Requests for additional information may be directed to the corresponding author and will be considered in accordance with applicable privacy regulations and organizational data-sharing policies.

## Acknowledgments

The authors acknowledge the Virta Health clinical care, health coaching, and data analytics teams for their contributions to the delivery of the obesity care program and the collection and maintenance of clinical data used in this study.

During the preparation of this manuscript, the authors used ChatGPT (OpenAI) to assist with language editing, grammatical refinement, and improving the clarity and organization of manuscript text. The authors reviewed and edited the output and take full responsibility for the content of this publication.

## Conflicts of Interest

S.J.A., A.R.Z., and K.E.B. are employees of Virta Health and hold stock or stock options in the company. A.M.G. and B.A.G. declare no conflicts of interest [subject to author confirmation]. Virta Health provided the clinical care infrastructure and data used in this study. The authors were responsible for the study design, data analysis, interpretation of findings, manuscript preparation, and decision to submit the manuscript for publication.

**Disclaimer/Publisher’s Note:** The statements, opinions and data contained in all publications are solely those of the individual author(s) and contributor(s) and not of MDPI and/or the editor(s). MDPI and/or the editor(s) disclaim responsibility for any injury to people or property resulting from any ideas, methods, instructions or products referred to in the content.

## Notes

### Competing Interest Statement

SJA, ARZ and KEB are employees of Virta Health and hold stock or stock options in the company.

### Author Declarations

This retrospective, real-world observational study utilized exclusively de-identified clinical data obtained from existing medical records. All patient information was de-identified in accordance with the Health Insurance Portability and Accountability Act (HIPAA) regulations to protect patient privacy and confidentiality. The study was exempt from formal ethical approval requirements because it involved secondary analysis of de-identified data without identifiable human subjects. The study was conducted in accordance with the principles of the Declaration of Helsinki and its subsequent amendments.

### Summary of Updates

This is the revised version after full review

