## Supplemental Material for "Real-World Twelve-Month Weight Loss Outcomes in a Comprehensive Obesity Care Program Utilizing Individualized Nutrition Therapy and Incretin-Based Obesity Medications"

**Supplementary Figure S1.** Longitudinal body weight trajectories through 15 months among GLP-1–naïve participants receiving semaglutide or tirzepatide by treatment pathway.

Mean body weight (kg) at program enrollment and 6, 12, and 15 months among GLP-1–naïve participants receiving a single GLP-1–based medication (semaglutide or tirzepatide) in the VINT+IOM and IOM only pathways. Data points represent mean body weight, with error bars indicating variability around the estimates. Numbers below each time point indicate the number of participants with available weight measurements. VINT, Virta Individualized Nutrition Therapy; IOM, incretin-based obesity medication.

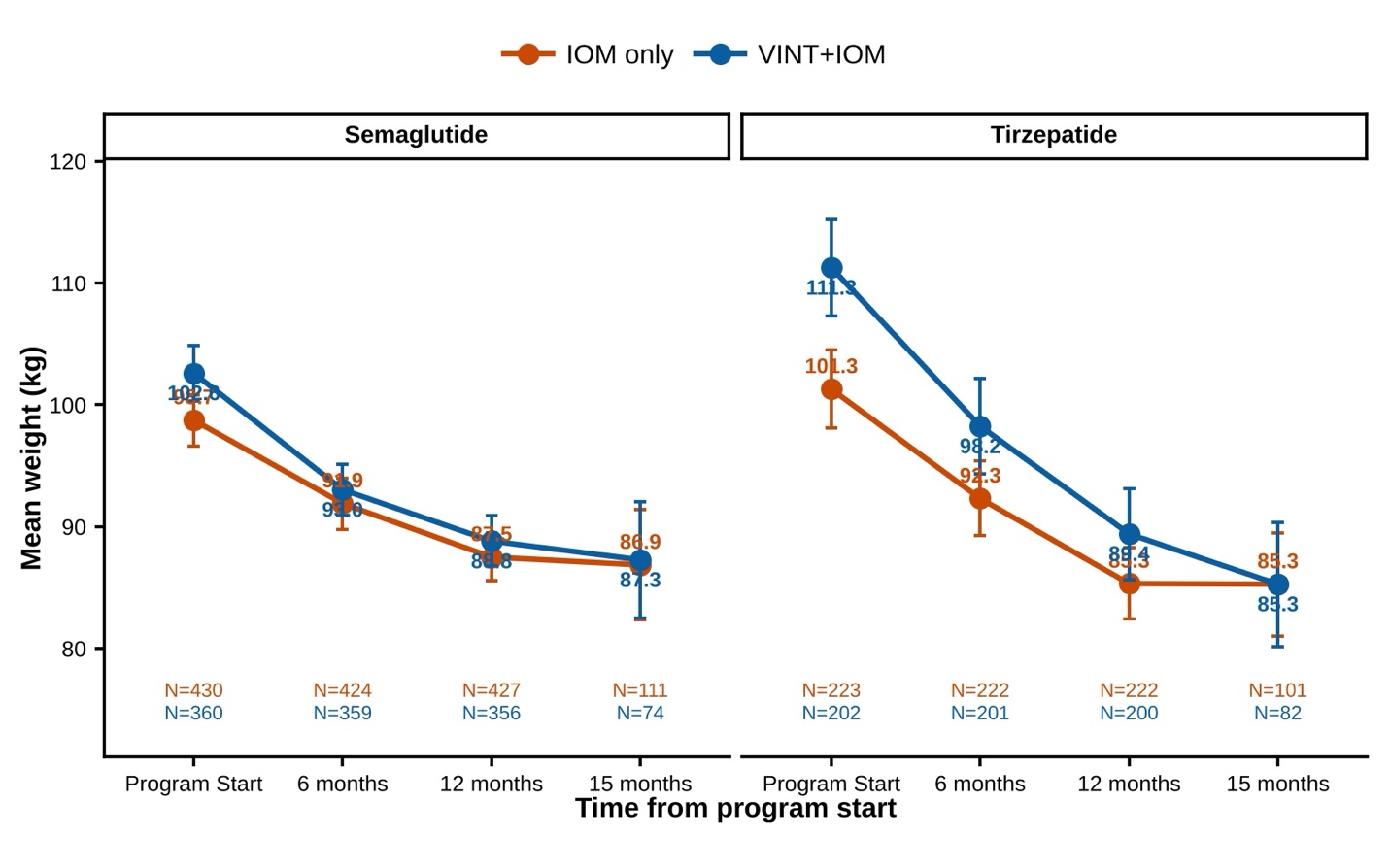

**Supplementary Figure S2.** Adjusted mean body weight by sex over 12 months among GLP-1-naïve participants receiving a single GLP-1-based ingredient. Panels show adjusted mean weight (kg) at program start, 6 months, and 12 months for women (purple) and men (green) in the following groups: (A) semaglutide, IOM only; (B) semaglutide, VINT+IOM; (C) tirzepatide, IOM only; (D) tirzepatide, VINT+IOM. Points are model-based estimated marginal means, and error bars show 95% confidence intervals. Estimates come from longitudinal linear mixed-effects models with a participant-level random intercept, including time × sex × pathway × medication terms and adjusted for age, baseline HbA1c, and baseline BMI. Values beside each point are adjusted means in lb. IOM, incretin-based obesity medication; VINT, Virta Individualized Nutrition Therapy.

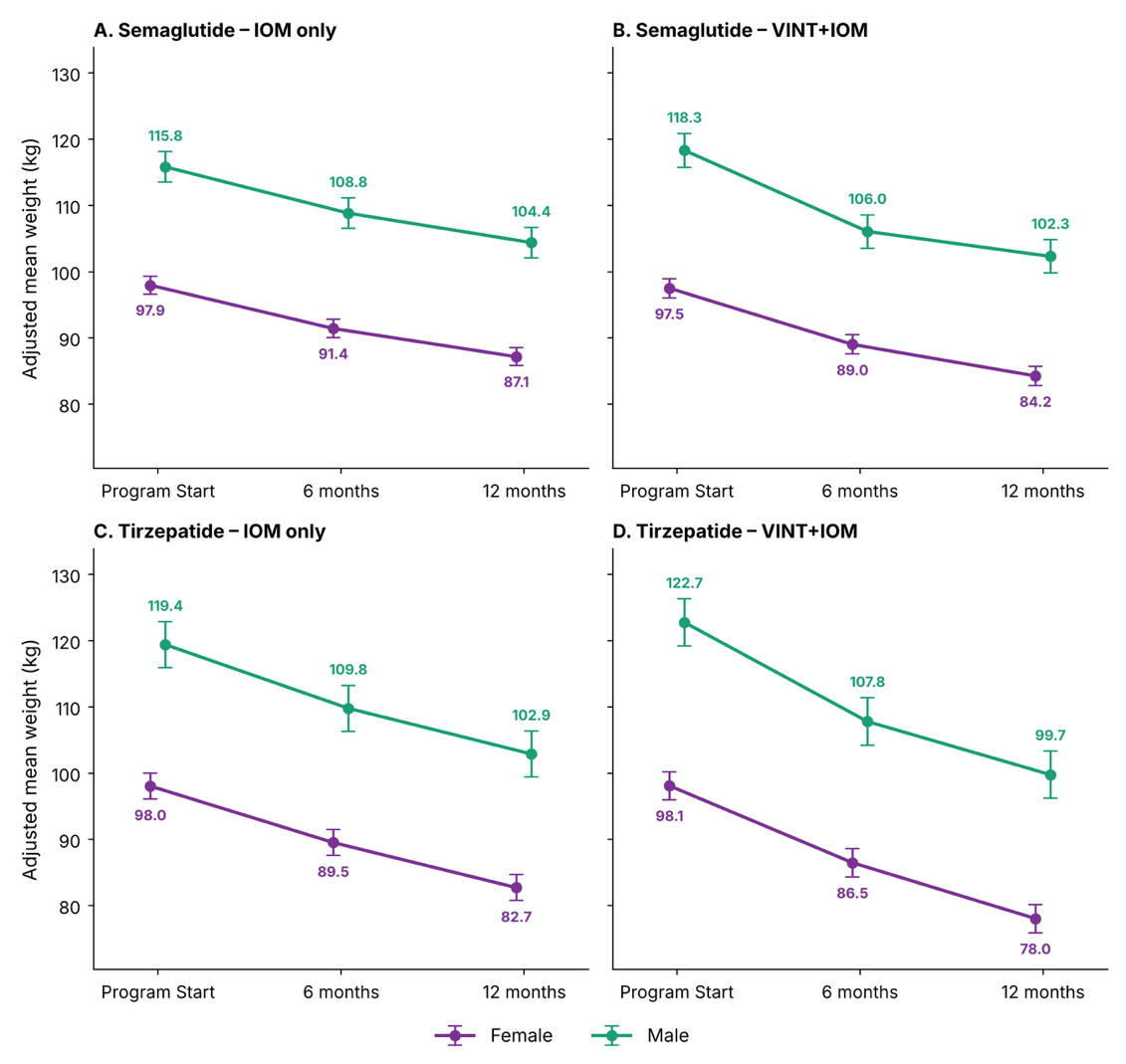

**Supplementary Tables**

**Supplementary Table S1.** Adjusted Estimated Marginal Mean Weight Outcomes Across Cohorts and Medication Subgroups

| **Pathway** | **Medication** | **Pre-GLP-1**  **EMM, lb (95% CI)** | **Program-start**  **EMM, lb (95% CI)** | **6-mo**  **EMM, lb (95% CI)** | **6-mo**  **change, lb** | **12-mo EMM, lb (95% CI)** | **12-mo**  **change,**  **lb** | **WG**  **P value** | **BG**  **P value** |
| --- | --- | --- | --- | --- | --- | --- | --- | --- | --- |
| **Primary outcomes** | | | | | | | | | |
| **Single ingredient (GLP-1 naive)** | | | | | | | | | |
| IOM | Semaglutide | — | 237.7 (223.4 to 252.0) | 223.1 (208.8 to 237.4) | −14.6 (−16.6 to −12.7) | 213.6 (199.3 to 227.8) | −24.1 (−26.1 to −22.2) | <0.001 | ref |
| IOM | Tirzepatide | — | 239.8 (225.1 to 254.5) | 220.4 (205.8 to 235.1) | −19.4 (−22.1 to −16.6) | 205.4 (190.7 to 220.0) | −34.4 (−37.2 to −31.7) | <0.001 | ref |
| VINT+IOM | Semaglutide | — | 238.3 (223.9 to 252.8) | 217.6 (203.1 to 232.0) | −20.8 (−22.8 to −18.7) | 207.6 (193.1 to 222.0) | −30.7* (−32.8 to −28.7) | <0.001 | 0.012 |
| VINT+IOM | Tirzepatide | — | 241.9 (227.1 to 256.7) | 214.4 (199.6 to 229.1) | −27.5 (−30.5 to −24.5) | 196.0 (181.2 to 210.7) | −45.9* (−48.9 to −42.9) | <0.001 | 0.003 |
| **Ancillary outcomes** | | | | | | | | | |
| **Overall Rx cohort** | | | | | | | | | |
| IOM | All | — | 226.2 (219.9 to 232.5) | 211.9 (205.6 to 218.2) | −14.3 (−15.0 to −13.5) | 204.7 (198.4 to 211.0) | −21.5 (−22.3 to −20.7) | <0.001 | ref |
| VINT+IOM | All | — | 226.3 (219.9 to 232.6) | 206.2 (199.9 to 212.6) | −20.1 (−21.0 to −19.1) | 196.9 (190.6 to 203.3) | −29.4* (−30.3 to −28.4) | <0.001 | <0.001 |
| **GLP-1 naive** | | | | | | | | | |
| IOM | All | — | 229.7 (219.3 to 240.0) | 212.8 (202.5 to 223.2) | −16.9 (−18.1 to −15.6) | 202.4 (192.1 to 212.8) | −27.2 (−28.5 to −26.0) | <0.001 | ref |
| VINT+IOM | All | — | 229.4 (219.0 to 239.9) | 206.0 (195.5 to 216.4) | −23.5 (−24.8 to −22.1) | 194.4 (183.9 to 204.8) | −35.1* (−36.4 to −33.7) | <0.001 | <0.001 |
| **Prior GLP-1** |  |  |  |  |  |  |  |  |  |
| IOM | All | 255.7 (247.6 to 263.8) | 223.0 (214.9 to 231.1) | 210.8 (202.7 to 218.9) | −12.2 (−13.5 to −11.0) | 206.1 (198.0 to 214.2) | −16.9 (−18.2 to −15.7) | <0.001 | ref |
| VINT+IOM | All | 248.9 (240.7 to 257.1) | 223.2 (215.0 to 231.4) | 207.2 (199.0 to 215.4) | −16.1 (−17.8 to −14.3) | 200.5 (192.3 to 208.7) | −22.7* (−24.4 to −21.0) | <0.001 | <0.001 |
| **Single ingredient (Prior GLP-1)** | | | | | | | | | |
| IOM | Semaglutide | 254.0 (245.8 to 262.2) | 220.0 (211.8 to 228.2) | 210.1 (201.9 to 218.2) | −9.9 (−11.8 to −8.1) | 206.8 (198.6 to 215.0) | −13.2 (−15.1 to −11.3) | <0.001 | ref |
| IOM | Tirzepatide | 253.1 (244.3 to 261.9) | 220.9 (212.2 to 229.7) | 202.8 (194.0 to 211.6) | −18.1 (−21.2 to −15.0) | 194.2 (185.4 to 203.0) | −26.8 (−29.8 to −23.7) | <0.001 | ref |
| VINT+IOM | Semaglutide | 246.5 (238.1 to 255.0) | 219.8 (211.4 to 228.3) | 205.7 (197.3 to 214.2) | −14.1 (−16.6 to −11.6) | 199.8 (191.3 to 208.2) | −20.1* (−22.6 to −17.6) | <0.001 | 0.018 |
| VINT+IOM | Tirzepatide | 244.9 (235.5 to 254.4) | 220.2 (210.8 to 229.6) | 199.5 (190.0 to 208.9) | −20.7 (−25.0 to −16.4) | 188.3 (178.8 to 197.7) | −31.9* (−36.2 to −27.6) | <0.001 | 0.035 |

***NOTE:*** *Values are adjusted estimated marginal means (EMMs) with 95% confidence intervals from longitudinal mixed-effects models. Negative changes indicate weight loss from program start. All program-start to 12-month within-group changes were statistically significant (P < 0.001).
 * Indicates a statistically significant between-group difference (VINT+IOM vs. IOM only within the respective cohort/medication subgroup, P < 0.05). 'ref' indicates the baseline reference group for between-group P-value comparisons.* ***ABBREVIATIONS****: VINT, Virta Individualized Nutrition Intervention; IOM, Incretin based obesity medication; mo, month; WG, within-group; BG, between-group*

**Supplementary Table S2.** Tests of sex-by-treatment interaction terms in the weight linear mixed models

| Term | F | df (num, den) | p-value |
| --- | --- | --- | --- |
| A. time × sex | | | |
| Sex | 648.12 | 1, 915 | **<0.001** |
| Time × Sex | 5.91 | 2, 1819 | **0.003** |
| B. time × sex × pathway | | | |
| Sex | 649.56 | 1, 915 | **<0.001** |
| Time × Sex | 6.59 | 2, 1819 | **0.001** |
| Sex × Pathway | 0.98 | 1, 914 | 0.322 |
| Time × Sex × Pathway | 3.24 | 2, 1819 | **0.039** |
| C. time × sex × medication | | | |
| Sex | 605.87 | 1, 914 | **<0.001** |
| Time × Sex | 6.04 | 2, 1819 | **0.002** |
| Sex × Medication | 4.93 | 1, 914 | **0.027** |
| Time × Sex × Medication | 0.12 | 2, 1819 | 0.891 |
| D. time × sex × pathway × medication | | | |
| Sex | 607.62 | 1, 914 | **<0.001** |
| Time × Sex | 6.37 | 2, 1819 | **0.002** |
| Sex × Pathway | 0.94 | 1, 914 | 0.331 |
| Sex × Medication | 4.90 | 1, 914 | **0.027** |
| Time × Sex × Pathway | 2.46 | 2, 1819 | 0.086 |
| Time × Sex × Medication | 0.05 | 2, 1819 | 0.953 |
| Sex × Pathway × Medication | 0.07 | 1, 914 | 0.798 |
| Time × Sex × Pathway × Medication | 0.09 | 2, 1819 | 0.917 |

*Notes: Linear mixed models of weight (lb) with a random intercept for participant, fit by maximum likelihood, including female and male participants only (n = 683 women and 231 men; 2,731 weight observations) and adjusted for age, baseline HbA1c and baseline BMI. Models A–D differ in which treatment factors enter the interaction with sex and time; only terms involving sex are shown. Pathway: IOM only vs VINT+IOM. Medication: semaglutide vs tirzepatide. Terms containing Time test whether weight change over time differs by sex (and by treatment); terms without Time test baseline-level differences. F tests use Satterthwaite degrees of freedom; p-values are not adjusted for multiplicity. Bold = p < 0.05.*

**Supplementary Table S3.** Observed and adjusted weight trajectories by sex, medication and pathway

|  |  | Observed weight, lb, mean (SD) | | | Adjusted weight, lb, mean (95% CI) | | |
| --- | --- | --- | --- | --- | --- | --- | --- |
| Sex | **N** | **Program Start** | **6 months** | **12 months** | **Program Start** | **6 months** | **12 months** |
| Semaglutide – IOM only | | | | | | | |
| Female | 248 | 206.9 (42.2) | 192.7 (40.3) | 183.1 (38.9) | 215.9 (212.9, 218.9) | 201.5 (198.5, 204.5) | 192.1 (189.1, 195.1) |
| Male | 86 | 249.1 (49.4) | 233.6 (49.0) | 223.8 (47.4) | 255.3 (250.2, 260.3) | 239.9 (234.8, 245.0) | 230.1 (225.0, 235.2) |
| Semaglutide – VINT+IOM | | | | | | | |
| Female | 211 | 216.4 (43.8) | 197.8 (41.0) | 187.4 (39.4) | 214.9 (211.6, 218.1) | 196.2 (193.0, 199.5) | 185.7 (182.4, 188.9) |
| Male | 71 | 264.2 (51.6) | 237.2 (45.3) | 228.9 (44.3) | 260.8 (255.2, 266.4) | 233.8 (228.2, 239.3) | 225.5 (219.9, 231.1) |
| Tirzepatide – IOM only | | | | | | | |
| Female | 122 | 213.7 (48.6) | 194.9 (46.5) | 179.9 (43.9) | 216.1 (211.8, 220.4) | 197.3 (193.0, 201.6) | 182.3 (178.0, 186.6) |
| Male | 38 | 269.3 (65.7) | 248.2 (59.7) | 232.9 (53.2) | 263.2 (255.6, 270.8) | 242.0 (234.4, 249.7) | 226.8 (219.1, 234.4) |
| Tirzepatide – VINT+IOM | | | | | | | |
| Female | 102 | 229.2 (53.1) | 204.2 (53.5) | 183.9 (48.6) | 216.2 (211.6, 220.9) | 190.6 (185.9, 195.3) | 172.0 (167.3, 176.7) |
| Male | 36 | 296.2 (76.4) | 263.2 (76.9) | 245.4 (75.2) | 270.6 (262.7, 278.5) | 237.6 (229.7, 245.5) | 219.9 (212.0, 227.7) |

*Notes: N = participants with a baseline weight in the sex model. Observed values are raw means among participants with complete model covariates (age, baseline HbA1c, baseline BMI), so they are a subset of the full cohort (914 of 1,214 women and men; the rest lack baseline HbA1c). Adjusted values are model-estimated means (time × sex × pathway × medication model) at the average age, baseline HbA1c and baseline BMI of the sample, so they differ from the observed means, especially at baseline.*

**Supplementary Table S4.** Adjusted change in weight from program start by sex, and sex difference in change

| Timepoint | Women, change in lb (95% CI) | Men, change in lb (95% CI) | Difference in change, women − men, lb (95% CI) | p-value |
| --- | --- | --- | --- | --- |
| Semaglutide – IOM only | | | | |
| 6 months | −14.4 (−16.6, −12.2) | −15.3 (−19.1, −11.5) | 0.9 (−3.5, 5.3) | 0.678 |
| 12 months | −23.9 (−26.1, −21.7) | −25.2 (−29.0, −21.4) | 1.3 (−3.1, 5.7) | 0.560 |
| Semaglutide – VINT+IOM | | | | |
| 6 months | −18.6 (−21.1, −16.2) | −27.0 (−31.2, −22.9) | 8.4 (3.6, 13.2) | **<0.001** |
| 12 months | −29.2 (−31.6, −26.8) | −35.3 (−39.5, −31.2) | 6.1 (1.3, 10.9) | **0.012** |
| Tirzepatide – IOM only | | | | |
| 6 months | −18.8 (−22.0, −15.6) | −21.1 (−26.8, −15.5) | 2.3 (−4.2, 8.8) | 0.483 |
| 12 months | −33.8 (−37.0, −30.6) | −36.4 (−42.1, −30.7) | 2.6 (−3.9, 9.1) | 0.433 |
| Tirzepatide – VINT+IOM | | | | |
| 6 months | −25.6 (−29.1, −22.1) | −33.0 (−38.8, −27.2) | 7.4 (0.6, 14.2) | **0.033** |
| 12 months | −44.2 (−47.7, −40.7) | −50.8 (−56.6, −44.9) | 6.6 (−0.2, 13.4) | 0.059 |

***Notes****: Estimates are from the time × sex × pathway × medication model (adjusted for age, baseline HbA1c and baseline BMI). Change = weight at the timepoint minus weight at program start, so negative values are weight loss. A positive difference (women − men) means women lost fewer pounds than men. p-values test the difference in change between sexes within each medication–pathway group and are not adjusted for multiplicity. Bold = p < 0.05. Within-sex changes from program start were significant (Bonferroni-adjusted p < 0.001)* *in every group.*

**Supplementary Table S5.** Observed percentage weight loss and responder thresholds at 12 months, by sex

| Sex | N | Weight loss, % (mean, SD) | ≥5% loss, n (%) | ≥10% loss, n (%) | ≥15% loss, n (%) | ≥20% loss, n (%) |
| --- | --- | --- | --- | --- | --- | --- |
| All participants | | | | | | |
| Female | 903 | 14.3 (9.7) | 759 (84.1) | 607 (67.2) | 425 (47.1) | 247 (27.4) |
| Male | 301 | 13.2 (10.2) | 247 (82.1) | 193 (64.1) | 124 (41.2) | 64 (21.3) |
| *p (women vs men)* |  | *—* | *0.420* | *0.325* | *0.082* | ***0.040*** |
| Semaglutide – IOM only | | | | | | |
| Female | 321 | 11.5 (9.2) | 242 (75.4) | 188 (58.6) | 118 (36.8) | 59 (18.4) |
| Male | 105 | 10.0 (10.6) | 76 (72.4) | 54 (51.4) | 31 (29.5) | 10 (9.5) |
| *p (women vs men)* |  | *—* | *0.605* | *0.213* | *0.196* | ***0.033*** |
| Semaglutide – VINT+IOM | | | | | | |
| Female | 263 | 13.4 (8.3) | 229 (87.1) | 170 (64.6) | 107 (40.7) | 54 (20.5) |
| Male | 93 | 13.0 (8.4) | 80 (86.0) | 60 (64.5) | 35 (37.6) | 16 (17.2) |
| *p (women vs men)* |  | *—* | *0.859* | *1.000* | *0.624* | *0.546* |
| Tirzepatide – IOM only | | | | | | |
| Female | 172 | 15.9 (10.5) | 145 (84.3) | 123 (71.5) | 99 (57.6) | 65 (37.8) |
| Male | 50 | 14.8 (10.4) | 41 (82.0) | 36 (72.0) | 24 (48.0) | 16 (32.0) |
| *p (women vs men)* |  | *—* | *0.668* | *1.000* | *0.260* | *0.507* |
| Tirzepatide – VINT+IOM | | | | | | |
| Female | 147 | 20.1 (9.1) | 143 (97.3) | 126 (85.7) | 101 (68.7) | 69 (46.9) |
| Male | 53 | 18.6 (9.6) | 50 (94.3) | 43 (81.1) | 34 (64.2) | 22 (41.5) |
| *p (women vs men)* |  | *—* | *0.385* | *0.507* | *0.609* | *0.524* |

*Notes: Observed data in all participants with a weight at 9–12 months (not restricted to participants with complete model covariates). Percentage weight loss = (baseline weight − follow-up weight) / baseline weight × 100. Responder percentages use participants with a follow-up weight as the denominator. p-values compare responder proportions between women and men (Fisher exact test), within each group; they are not adjusted for multiplicity (20 tests in this table). Bold = p<0.05*

**Supplementary Table S6.** Total Weight Loss from the Pre-GLP-1 Reference Among Participants with Prior GLP-1 Exposure

| Pathway | Medication | Cohort N | Mean % loss, pre-GLP-1 to program start | Mean % loss, pre-GLP-1 to 6 mo | Mean % loss, pre-GLP-1 to 12 mo | 12-mo n | ≥5% weight loss, n (%) | ≥10% weight loss, n (%) | ≥15% weight loss, n (%) | ≥20% weight loss, n (%) |
| --- | --- | --- | --- | --- | --- | --- | --- | --- | --- | --- |
| Prior GLP-1 | | | | | | | | | | |
| Lifestyle+IOM | All medications | 1,482 | 13.4% | 18.5% | 20.4% | 1,464 | 1,408 (96.2%) | 1,296 (88.5%) | 1,018 (69.5%) | 705 (48.2%) |
| VINT+IOM | All medications | 763 | 10.6% | 17.1% | 19.7% | 761 | 737 (96.8%) | 653 (85.8%) | 509 (66.9%) | 348 (45.7%) |
| Single ingredient (Prior GLP-1) | | | | | | | | | | |
| Lifestyle+IOM | Semaglutide | 677 | 14.1% | 18.5% | 19.8% | 669 | 637 (95.2%) | 587 (87.7%) | 457 (68.3%) | 310 (46.3%) |
| Lifestyle+IOM | Tirzepatide | 240 | 13.4% | 21.0% | 24.5% | 240 | 239 (99.6%) | 229 (95.4%) | 197 (82.1%) | 154 (64.2%) |
| VINT+IOM | Semaglutide | 349 | 11.0% | 17.0% | 19.2% | 349 | 337 (96.6%) | 292 (83.7%) | 231 (66.2%) | 149 (42.7%) |
| VINT+IOM | Tirzepatide | 120 | 10.2% | 18.5% | 23.0% | 119 | 119 (100.0%) | 116 (97.5%) | 91 (76.5%) | 71 (59.7%) |

*These outcomes use the pre-GLP-1 weight as the reference and therefore represent total weight loss accrued before and after entry into the SWL prescription pathway. Responder percentages are calculated among participants with an available weight at 9–12 months.*

**Supplementary Table S7.** Medication utilization by Rx pathways

Values are n/N (%) unless otherwise specified. Count outcomes are median (Q1–Q3). P values are unadjusted.

| Outcome | IOM only (n=2,611) | VINT+IOM(n=1,658) | P value |
| --- | --- | --- | --- |
| Starting medication: semaglutide | 1,346/2,611 (51.6%) | 832/1,658(50.2%) | 0.639 |
| Starting medication: tirzepatide | 1,256/2,611(48.1%) | 819/1,658(49.4%) |  |
| Starting medication: liraglutide | 9/2,611(0.9%) | 7/1,658(0.4%) |  |
| Active on GLP-1 at 12 months | 2,163/2,611 (82.8%) | 1,326/1,658 (80.0%) | 0.018 |
| Switched GLP-1 ingredient | 1,028/2,611 (39.4%) | 623/1,658 (37.6%) | 0.240 |
| Final medication: semaglutide | 1,873/2,611 (71.7%) | 1,166/1,658 (70.3%) | 0.567 |
| Final medication: tirzepatide | 732/2,611 (28.0%) | 487/1,658 (29.4%) |  |
| Final medication: liraglutide | 6/2,611 (0.2%) | 5/1,658 (0.3%) |  |
| Number of distinct ingredients | 1 (1–2) | 1 (1–2) | 0.261 |
| *Overall comparison across final-medication categories; interpret cautiously because liraglutide use was rare. | | | |

**Note.** Values are presented as n/N (%) unless otherwise specified; count outcomes are presented as median (Q1–Q3). P values are unadjusted. Categorical outcomes were compared between pathways using chi-square tests; count outcomes were compared using Wilcoxon rank-sum tests. The P value for final medication represents an overall comparison across medication-ingredient categories and should be interpreted cautiously because liraglutide use was rare. IOM, incretin-based obesity medication; Q1, first quartile; Q3, third quartile; VINT, Virta individualized nutrition therapy.

**Supplementary Table S8.** Medication trajectories for 12 months

Trajectory categories were updated using participant-level validation of apparent discontinuations. The overall distribution differed between pathways (chi-square=50.34; df=7; P<0.001).

| Medication trajectory | IOM only n (%) | VINT + IOM n (%) |
| --- | --- | --- |
| De-escalation only | 96 (3.7%) | 38 (2.3%) |
| Discontinued and restarted | 355 (13.6%) | 222 (13.4%) |
| Discontinued before 12 months | 42 (1.6%) | 28 (1.7%) |
| Escalation and de-escalation | 428 (16.4%) | 288 (17.4%) |
| Escalation only | 546 (20.9%) | 460 (27.7%) |
| Ingredient switch | 762 (29.2%) | 452 (27.3%) |
| Ingredient switch followed by discontinuation | 78 (3.0%) | 53 (3.2%) |
| Stable/other | 304 (11.6%) | 117 (7.1%) |

Validation rule: participants with no documented restart through the observation end remained in the corresponding discontinuation category. Those restarting after 31-90 days or after >90 days were reclassified as discontinued and restarted. Records showing resumed or continued therapy within 30 days were not considered confirmed discontinuations; these participants were reclassified as ingredient switch when an ingredient switch was documented, or stable/other otherwise. Percentages use pathway-specific denominators of 2,611 and 1,658.

**Note.** Values are presented as n (%), using pathway-specific denominators of 2,611 and 1,658. Trajectory categories were updated following participant-level validation of apparent discontinuations. Participants with no documented restart through the observation period remained classified in the corresponding discontinuation category. Participants with a documented restart after 31–90 days or >90 days were classified as discontinued and restarted, whereas resumed or continued therapy within 30 days was not considered a confirmed discontinuation; these participants were classified as ingredient switch when an ingredient switch was documented, or stable/other otherwise. The overall distribution of medication trajectories was compared between pathways using a chi-square test. IOM, incretin-based obesity medication; VINT, Virta individualized nutrition therapy.

The overall distribution of medication trajectories differed between pathways (χ²=50.34; df=7; *P*<0.001).

**Supplementary Table S9.** Overall medication titration by Rx pathway

Binary outcomes are n/N (%); count and time outcomes are median (Q1–Q3). Days to maximum dose are restricted to 0–366 days.

| Outcome | IOM only (n=2,611) | VINT+IOM (n=1,658) | P value |
| --- | --- | --- | --- |
| Ever dose escalation | 2,030/2,611 (77.7%) | 1,406/1,658 (84.8%) | <0.001 |
| Ever dose de-escalation | 977/2,611 (37.4%) | 632/1,658 (38.1%) | 0.646 |
| Number of dose increases | 2.0 (1.0–3.0) | 3.0 (1.0–4.0) | <0.001 |
| Number of dose decreases | 0.0 (0.0–1.0) | 0.0 (0.0–1.0) | 0.752 |
| Days to maximum dose | 76.0 (0.0–154.2) | 99.0 (17.0–163.0) | <0.001 |

**Note.** Binary outcomes are presented as n/N (%); count and time outcomes are presented as median (Q1–Q3). Dose escalation and de-escalation were determined from sequential dose changes within the same medication ingredient when dose units and administration frequencies were comparable. Days to maximum dose was restricted to values from 0 to 366 days. Binary outcomes were compared between pathways using chi-square tests, and count and time outcomes were compared using Wilcoxon rank-sum tests. IOM, incretin-based obesity medication; Q1, first quartile; Q3, third quartile; VINT, Virta individualized nutrition therapy.

**Supplementary Table S10.** Ingredient-specific medication doses by Rx pathway

Analyses use clinically plausible dose ranges (semaglutide ≤2.4 mg; tirzepatide ≤15 mg). Semaglutide and tirzepatide doses were analyzed separately.

| Dose stage | Ingredient | IOM only, median (Q1–Q3) | n | VINT+IOM, median (Q1–Q3) | n | P value |
| --- | --- | --- | --- | --- | --- | --- |
| Starting dose | Semaglutide | 1.7 (0.5–2.4) | 1346 | 0.5 (0.3–2.4) | 832 | <0.001 |
| Starting dose | Tirzepatide | 5.0 (2.5–10.0) | 1256 | 5.0 (2.5–7.5) | 819 | <0.001 |
| Final dose | Semaglutide | 2.4 (1.7–2.4) | 1873 | 2.4 (1.7–2.4) | 1166 | 0.344 |
| Final dose | Tirzepatide | 12.5 (7.5–15.0) | 732 | 12.5 (7.5–15.0) | 487 | 0.856 |

**Note.** Values are presented as median (Q1–Q3), with the number of participants contributing dose data (n) shown separately. Starting dose was defined as the dose active at program start or, if no medication was active at program start, the first dose initiated thereafter. Final dose was defined as the dose active at 12 months or, if no medication was active at 12 months, the last observed dose before 12 months. Analyses were restricted to clinically plausible dose ranges for semaglutide (≤2.4 mg) and tirzepatide (≤15 mg), and ingredients were analyzed separately. P values are from Wilcoxon rank-sum tests comparing dose distributions between treatment pathways. IOM, individualized obesity medication; Q1, first quartile; Q3, third quartile; VINT, Virta individualized nutrition therapy.

**Supplementary Table S11. GLP-1 medication coverage by medication ingredient and Rx pathway among participants who stayed in a single ingredient**

| Medication ingredient | PDC measure | IOM only | VINT + IOM | P value |
| --- | --- | --- | --- | --- |
| Semaglutide | N | 1,116 | 711 |  |
|  | Mean PDC, % (SD) | 88.2 (16.9) | 89.2 (16.6) |  |
|  | Median PDC, % [IQR] | 95.6 [82.5, 99.7] | 98.1 [84.2, 100.0] | <0.001¹ |
|  | PDC <50%, n (%) | 47 (4.2) | 30 (4.2) |  |
|  | PDC 50% to <80%, n (%) | 196 (17.6) | 113 (15.9) |  |
|  | PDC 80% to <90%, n (%) | 163 (14.6) | 105 (14.8) |  |
|  | PDC ≥90%, n (%) | 710 (63.6) | 463 (65.1) |  |
|  | PDC ≥80%, n (%) | 873 (78.2) | 568 (79.9) | 0.396² |
| Tirzepatide | N | 463 | 322 |  |
|  | Mean PDC, % (SD) | 86.9 (19.0) | 87.1 (19.3) |  |
|  | Median PDC, % [IQR] | 94.8 [83.3, 99.5] | 95.9 [82.8, 99.5] | 0.637¹ |
|  | PDC <50%, n (%) | 37 (8.0) | 21 (6.5) |  |
|  | PDC 50% to <80%, n (%) | 55 (11.9) | 44 (13.7) |  |
|  | PDC 80% to <90%, n (%) | 89 (19.2) | 49 (15.2) |  |
|  | PDC ≥90%, n (%) | 282 (60.9) | 208 (64.6) |  |
|  | PDC ≥80%, n (%) | 371 (80.1) | 257 (79.8) | 0.913² |

Data are presented as mean (SD), median [IQR], or n (%). PDC was available for all 2,601 participants included in this analysis. Percentages are calculated within each medication ingredient and pathway.

¹ Wilcoxon rank-sum test comparing continuous PDC distributions between pathways.

² Pearson chi-square test comparing PDC ≥80% versus <80% between pathways.

GLP-1, glucagon-like peptide-1; IOM, incretin-based obesity medication; IQR, interquartile range; PDC, proportion of days covered; SD, standard deviation; SWL, Sustainable Weight Loss; VINT, Virta Individualized Nutrition Therapy.

**Supplementary Table S12. Observed 12-Month Weight Loss by GLP-1 Ingredient-Switching Direction, PDC Category, and SWL Pathway**

| Analysis subgroup | Pathway | Cohort N | 12 mo n | Mean (SD) | Median (IQR) | Pathway P value |
| --- | --- | --- | --- | --- | --- | --- |
| Panel A. Absolute weight loss, lb | | | | | | |
| Ingredient-switching direction | | | | | | |
| Semaglutide to tirzepatide | IOM only | 230 | 227 | 15.5 (21.4) | 12.3 (1.9–27.1) | 0.083 |
|  | VINT+IOM | 121 | 119 | 19.7 (21.4) | 17.0 (6.1–34.5) |  |
| Tirzepatide to semaglutide | Lifestyle+IOM | 788 | 772 | 23.2 (22.3) | 21.8 (8.5–35.6) | <0.001 |
|  | VINT+IOM | 494 | 491 | 30.8 (24.9) | 29.7 (13.4–45.9) |  |
| Medication adherence subgroup (PDC <80%) | | | | | | |
| PDC <50% | Lifestyle+IOM | 128 | 125 | 20.2 (22.8) | 19.1 (3.7–32.6) | 0.035 |
|  | VINT+IOM | 65 | 65 | 27.8 (23.7) | 24.8 (9.7–43.8) |  |
| PDC 50% to <80% | Lifestyle+IOM | 338 | 333 | 22.0 (23.9) | 18.1 (6.9–34.1) | <0.001 |
|  | VINT+IOM | 191 | 187 | 28.5 (24.1) | 28.3 (9.7–44.9) |  |

| Analysis subgroup | Pathway | Cohort N | 12 mo n | Mean (SD) | Median (IQR) | Pathway P value |
| --- | --- | --- | --- | --- | --- | --- |
| Panel B. Percentage weight loss, % | | | | | | |
| Ingredient-switching direction | | | | | | |
| Semaglutide to tirzepatide | Lifestyle+IOM | 230 | 227 | 6.5 (9.1) | 6.1 (1.0–11.6) | 0.042 |
|  | VINT+IOM | 121 | 119 | 8.5 (9.6) | 8.5 (2.3–13.4) |  |
| Tirzepatide to semaglutide | Lifestyle+IOM | 788 | 772 | 10.1 (9.2) | 9.9 (4.3–16.2) | <0.001 |
|  | VINT+IOM | 494 | 491 | 13.0 (10.1) | 13.3 (6.1–20.0) |  |
| Medication adherence subgroup (PDC <80%) | | | | | | |
| PDC <50% | Lifestyle+IOM | 128 | 125 | 9.2 (10.2) | 9.2 (1.7–15.6) | 0.077 |
|  | VINT+IOM | 65 | 65 | 11.9 (9.7) | 10.3 (5.7–18.4) |  |
| PDC 50% to <80% | Lifestyle+IOM | 338 | 333 | 9.7 (9.9) | 8.9 (3.1–15.5) | 0.004 |
|  | VINT+IOM | 191 | 187 | 11.9 (9.8) | 11.8 (4.7–19.7) |  |

Note. Switching direction was defined by the first and last documented GLP-1 ingredients during the 12-month observation period; intervening medication records may have occurred. PDC analyses included participants with observed-window PDC <80% (<50% or 50% to <80%). Weight loss was calculated from program enrollment to the available 12-month weight. Values are mean (SD) or median (IQR). Pathways were compared using the Wilcoxon rank-sum test; P values are unadjusted. Analyses were exploratory and unadjusted for baseline or treatment differences.

**Supplementary Table S13. Reported medication side effects during 12 months, by treatment pathway**

| Reported side effect or reporting measure | IOM only (n=2,611) | VINT+IOM (n=1,658) | P value |
| --- | --- | --- | --- |
| Any reported side effect, n (%) | 1,660 (63.6) | 1,071 (64.6) | 0.499 |
| Nausea, n (%) | 1,101 (42.2) | 695 (41.9) | 0.872 |
| Constipation, n (%) | 857 (32.8) | 616 (37.2) | 0.004 |
| Fatigue, n (%) | 736 (28.2) | 509 (30.7) | 0.079 |
| Diarrhea, n (%) | 480 (18.4) | 312 (18.8) | 0.722 |
| Vomiting, n (%) | 207 (7.9) | 107 (6.5) | 0.072 |
| Abdominal pain, n (%) | 0 (0.0) | 0 (0.0) | — |
| Days with a side-effect report, median (IQR)* | 4 (2–8) | 4 (2–8) | 0.403 |
| Total symptom reports, median (IQR)* | 5 (2–10) | 5 (2–10) | 0.377 |

**Note.** Side-effect records were linked to the analytic cohort by participant identifier and restricted to reports from program enrollment through 365 days. Values are participant-level counts and percentages unless otherwise specified.

Side effects reflect documented reports in longitudinal clinical logging data and should not be interpreted as systematically ascertained adverse-event incidence. Absence of a report was treated as no documented side effect.

***** Median and IQR are reported among participants with at least one documented side effect. P values for categorical outcomes were calculated using chi-square tests; continuous reporting measures were compared using Wilcoxon rank-sum tests.

No abdominal-pain reports were present in the source dataset.

**Supplementary Table S14.** Adjusted Estimated Marginal Mean Hemoglobin A1c Outcomes by Primary Rx Pathway Cohorts (Single Naïve GLP1 cohort) and Medication Subgroups

| Timepoint | N with HbA1c (% of group) | Adjusted mean HbA1c, % (95% CI) | Change from baseline, % points (95% CI) | p-value |
| --- | --- | --- | --- | --- |
| Semaglutide – IOM only (n = 430) | | | | |
| Program Start | 335 (78%) | 5.49 (5.29, 5.69) | — | — |
| 6 months | 136 (32%) | 5.28 (5.08, 5.48) | −0.21 (−0.26, −0.16) | <0.001 |
| 12 months | 166 (39%) | 5.22 (5.02, 5.42) | −0.27 (−0.31, −0.22) | <0.001 |
| Semaglutide – VINT+IOM (n = 360) | | | | |
| Program Start | 282 (78%) | 5.49 (5.29, 5.69) | — | — |
| 6 months | 115 (32%) | 5.29 (5.08, 5.49) | −0.20 (−0.26, −0.15) | <0.001 |
| 12 months | 138 (38%) | 5.19 (4.98, 5.39) | −0.31 (−0.36, −0.25) | <0.001 |
| Tirzepatide – IOM only (n = 223) | | | | |
| Program Start | 160 (72%) | 5.39 (5.19, 5.59) | — | — |
| 6 months | 76 (34%) | 5.16 (4.95, 5.36) | −0.23 (−0.30, −0.17) | <0.001 |
| 12 months | 94 (42%) | 5.03 (4.82, 5.24) | −0.36 (−0.42, −0.30) | <0.001 |
| Tirzepatide – VINT+IOM (n = 202) | | | | |
| Program Start | 138 (68%) | 5.53 (5.32, 5.73) | — | — |
| 6 months | 68 (34%) | 5.24 (5.03, 5.45) | −0.29 (−0.36, −0.22) | <0.001 |
| 12 months | 81 (40%) | 5.06 (4.85, 5.26) | −0.47 (−0.54, −0.40) | <0.001 |

*Notes: Values are model-based estimates from a linear mixed model with a random intercept for participant and a time × pathway × medication interaction, adjusted for age, gender and baseline BMI (1,789 HbA1c observations from 1,075 participants). Baseline HbA1c is the time-0 observation and is not used as a covariate. N is the number of participants with an HbA1c value at that timepoint; the percentage uses all participants in the group (n in the group header) as the denominator. The 6-month value is the 3-month result, or the 6-month result if the 6-month result was missing; the 12-month value is the 9-month result, or the 1-year result if the 1-year result was missing. Change from baseline = later timepoint minus baseline (negative = reduction); p-values are Bonferroni-adjusted for the three timepoint comparisons within each group, and confidence intervals are unadjusted.*
